# Pre-existing antibodies predict protection while mucosal inflammation correlates with symptomatic *Bordetella pertussis* infection

**DOI:** 10.64898/2026.06.26.26356067

**Authors:** Lisa Willemsen, Ziyuan Ren, Pramod Shinde, Nicola Thrupp, Jiyeun Lee, Aanya Gupta, Aaron Sutherland, Shelby Orfield, Mari Kojima, Ahmed Azhan, Jian Sun, April Frazier, Susan Hariri, Scott Halperin, May ElSherif, Bjoern Peters

**Affiliations:** Center for Infectious Disease and Vaccine Research, La Jolla Institute for Immunology, La Jolla, CA, USA; Division of Bacterial Diseases, National Center for Immunization and Respiratory Diseases, Centers for Disease Control and Prevention, 1600 Clifton Road NE, Atlanta, GA 30329, USA; Canadian Center for Vaccinology, Dalhousie University and the IWK Health Centre and Nova Scotia Health Authority, Halifax, Canada; Department of Medicine, University of California San Diego, La Jolla, CA, USA

## Abstract

Despite decades of widespread vaccination, whooping cough caused by *Bordetella pertussis* (Bp) continues to circulate globally. To define correlates of protection and mechanisms underlying symptom development, we characterized systemic and mucosal immune responses in a controlled human infection model of Bp (NCT05136599). Healthy vaccinated adults were intranasally challenged with escalating Bp doses and classified as symptomatic, asymptomatic, or non-infected. Longitudinal sampling of blood and nasal mucosa allowed mapping of antibody titers, immune cell subset frequencies, cytokine concentrations, gene expression, and T cell activation and polarization.

Non-infected participants exhibited significantly higher pre-challenge serum antigen-specific IgG titers. Post-challenge, only symptomatic participants developed robust antigen-specific IgG responses. Notably, infection-induced IgG responses displayed slower kinetics and a lower overall magnitude compared to the rapid day 7 peak observed following tetanus, diphtheria, and acellular pertussis (Tdap) booster vaccination. Furthermore, symptomatic infection was driven by pronounced nasal inflammation characterized by increased HLA-DRLJ myeloid cells and upregulated mucosal NF-κB signaling on day 7. Systemic immune responses were comparatively modest post-challenge: plasma cytokine concentrations decreased independently of clinical outcome, peripheral blood mononuclear cell transcriptomes and antigen-specific T cell activation and polarization remained largely unchanged.

These findings identify pre-existing serum antibodies as potential correlates of protection from Bp infection and suggest that symptom development is associated with localized mucosal inflammation dominated by a myeloid cell response. The predominance of nasal over systemic immune activation highlights the importance of mucosal immunity in controlling Bp and provides critical insights to guide the design of next-generation vaccines aimed at preventing both disease and transmission.

## Introduction

Whooping cough, caused by the highly contagious respiratory bacterium *Bordetella pertussis* (Bp), remains a significant global public health concern, marked by cyclical peaks in incidence and severe complications in young infants(1, 2). The introduction of the whole-cell Bp (wP) vaccine dramatically reduced disease incidence worldwide. However, due to high reactogenicity and public concern over largely unsubstantiated severe adverse events, many high-income countries replaced the wP vaccine with acellular pertussis (aP) vaccines. aP vaccines contain purified Bp antigens, including inactivated pertussis toxin (PT), filamentous hemagglutinin (FHA), pertactin (PRN), and in some formulations, fimbriae 2 and 3 (FIM), and are typically administered with tetanus and diphtheria toxoids(3). Although aP vaccines are associated with fewer side effects and effectively protect against severe symptoms, they confer less durable immunity and are less effective at preventing Bp infection and transmission compared to natural infection or wP vaccines(3–8). The recent resurgence of whooping cough, despite widespread vaccination coverage, underscores the need for improved vaccines that elicit more durable immunity and protection against infection. Therefore, a deeper understanding of protective immune responses, bacterial clearance, and symptom development is needed to inform future vaccine design.

Innate and adaptive immunity are critical for the control of Bp infections. Early innate responses, driven by myeloid cells and local inflammatory mediators, shape subsequent adaptive immunity and influence disease outcome(9). Natural infection, wP vaccination, and aP vaccination generate distinct adaptive immune responses with varying durability. Broad antigenic exposure from natural infection and wP vaccination drives a wider B and T cell repertoire, whereas aP vaccination elicits a narrower profile(10). Furthermore, natural infection and wP vaccination elicit more durable Th1 and Th17 responses, whereas aP vaccination typically induces a Th2-skewed response(4, 5, 11, 12). These T cell polarization phenotypes influence protection, with Th1/Th17 responses linked to more effective bacterial clearance(4, 11).

While these immune processes originate at the nasal mucosa—the primary site of Bp entry, infection, and transmission—they can also manifest systemically. To date, most human studies have focused on systemic immune responses due to the relative ease of blood sampling and the extensive standardization of blood-based assays. However, the nasal mucosa is equipped with diverse specialized immune mediators, including tissue-resident memory T cells, mucosal antibodies, and rapid-acting innate effectors. Higher bacterial loads at this site are known to trigger stronger innate and adaptive immune responses, influencing symptom severity and the development of durable immunity(13, 14). Yet, our understanding of nasal immunity to Bp in humans remains limited, hindered by challenges of mucosal sampling and the lack of well-timed exposure studies. In particular, the dynamic interplay between innate and adaptive immune responses in the nasal mucosa, and how these differ among symptomatic, asymptomatic, and resistant individuals, is poorly understood and may not be reflected in the systemic response.

Controlled human infection model (CHIM) studies provide a unique opportunity to study immune responses with precise timing of pathogen exposure and longitudinal sampling from multiple compartments. Combining serial blood and nasal sampling enables the tracking of immune activation kinetics, identification of correlates of protection, and dissection of cellular and molecular events that differentiate symptomatic, asymptomatic, and non-infected participants. These correlates could serve as candidate therapeutic targets, inform the development of next-generation pertussis vaccines, and streamline the evaluation of vaccine candidates without the need for full-scale clinical trials.

Here, we analyzed samples collected in a North American Bp CHIM study in which healthy adults were exposed to different bacterial doses(15). Using paired blood and nasal samples across multiple timepoints, we profiled gene expression in the nasal mucosa and blood, blood immune cell and cytokine composition, antigen-specific IgG responses, and T cell activation and polarization. This approach revealed both shared immune responses and responses specific to participant outcome (symptomatic, asymptomatic, or non-infected), with certain predictive immune profiles already detectable at baseline. We also observed substantial differences in immune responses induced by Bp challenge compared to tetanus, diphtheria, and acellular pertussis (Tdap) booster vaccination. These findings identify correlates of protection and provide mechanistic insight to help guide the development of next-generation pertussis vaccines.

## Results

### Profiling immune responses to Bp exposure via longitudinal blood and nasal sampling

All samples and participants described here were a subset of a recently published controlled human challenge model (NCT05136599)(15). Briefly, our cohort included healthy adults (n=53), aged 18-40 years, primed during infancy with either DTwP or DTaP vaccines (**Figure 1A, Table S1-3**). On day 0, participants were intranasally challenged with Bp (US Bp D420 strain) across four distinct dose groups ranging from 5x10L to 1×10L colony-forming units (CFU). Due to sample availability, participant numbers vary across individual assays; specific sample sizes for each experiment are detailed in **Table S3** and indicated within the respective figure legends.

**Figure 1.**
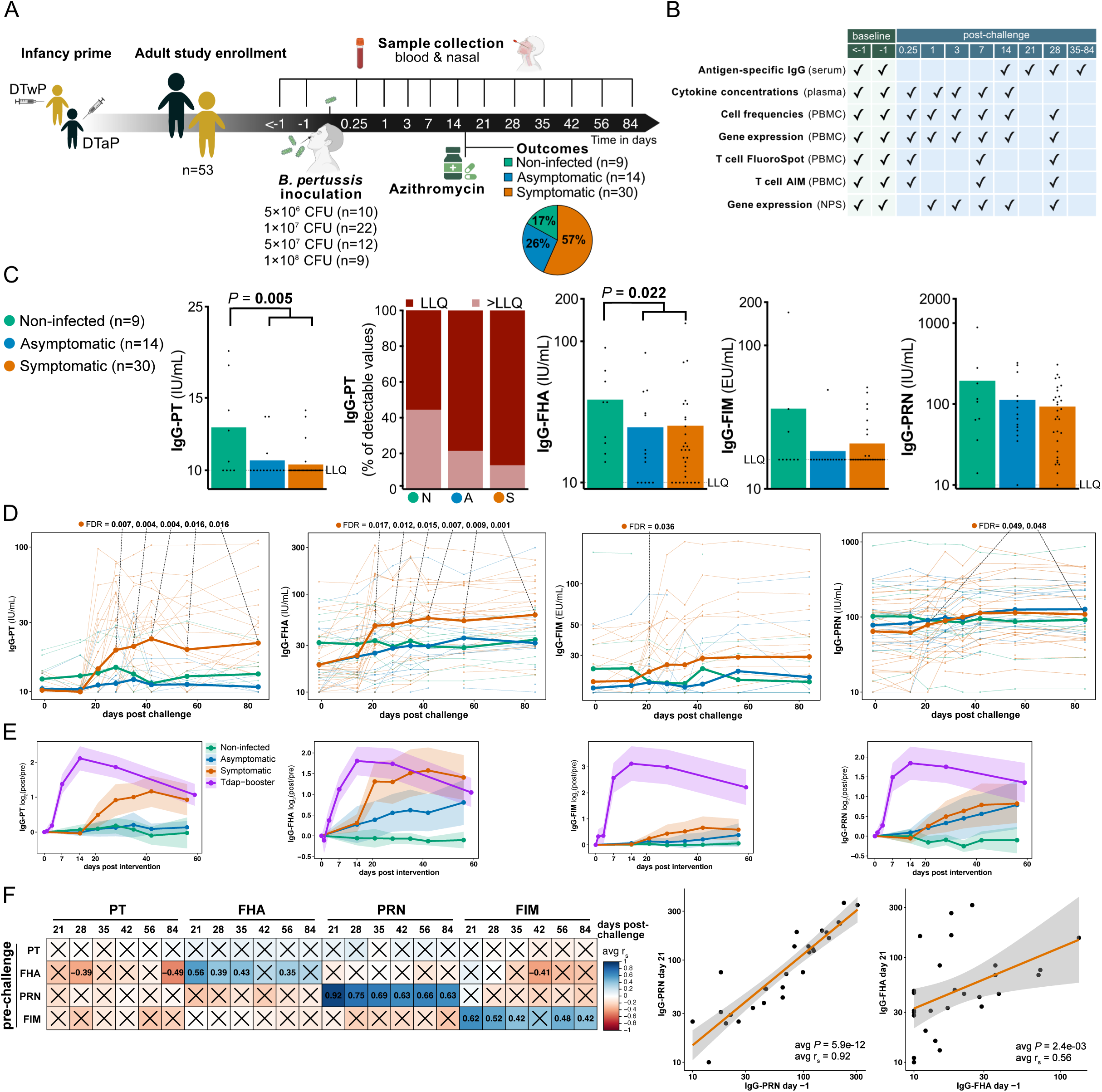
Non-infected participants had higher baseline Bp-specific serum IgG titers and Bp inoculation induced a humoral response in symptomatic participants in an antigen-specific fashion. **A)** Controlled Human Infection Model (CHIM) for Bordetella Pertussis (Bp) study outline. Healthy adults (n = 53) were recruited and primed with the DTaP or DTwP vaccine during infancy. Blood and nasal samples were collected before and after nasal inoculation with different doses of Bp. Participants were treated with azithromycin for 5 days. **B)** Performed experiments per timepoint. Serum and plasma were used for antibody and cytokine measurements respectively. Peripheral blood mononuclear cells (PBMC) were used to determine cell frequencies, gene expression, and T cell activation and polarization. Nasopharyngeal swabs (NPS) were used for gene expression and nasal washes (NW) and pharyngeal aspirates (NPA) for diagnostic purposes (not shown). **C-D)** Serum IgG titers against Bp antigens (PT, PRN, FHA, FIM) before (C-D) and after (D) Bp inoculation per outcome definition. LLQ, lower limit of quantification. Mean bars (C) and lines (D) are depicted per outcome definition. FDR values were calculated by performing (C) Mann-Whitney tests for two group comparisons, Kruskal-Wallis tests for three group comparisons followed by Dunn’s multiple comparisons test and (D) Dunnett’s multiple comparisons test for pairwise comparisons of post- versus pre-challenge timepoints per outcome using a mixed-effects model. **E)** Antigen-specific IgG responses (log_2_ post/pre) after Tdap booster vaccination (previous study) and Bp CHIM with mean lines depicted and 95% confidence interval shaded ribbons. **F)** Heatmap depicting the average Spearman (two-sided) correlation coefficients (rs) of serum IgG titers against Bp antigens (PT, PRN, FHA, FIM) of symptomatic participants before (rows) and after (columns) Bp challenge. Insignificant correlates are crossed (avg P > 0.1). Two correlations are shown: pre-challenge IgG-PRN versus IgG-PRN at day 21 (left) and pre-challenge IgG-FHA versus IgG-FHA at day 21 (right). The shaded area represents the 95% confidence interval around the regression line. Panels A-B were created using BioRender [license links].

Longitudinal blood and nasal samples were collected before and up to 84 days after Bp-challenge (**Figure 1B**). Blood samples were processed to obtain serum for antigen-specific antibody titer quantification, plasma for cytokine concentration measurements, and peripheral blood mononuclear cells (PBMC) for immune cell frequency profiling, gene expression analysis, and T cell activation and polarization studies. Nasal sampling included nasal washes (NW) and nasopharyngeal aspirates (NPA), both used for Bp detection via PCR and culture(15), and nasopharyngeal swabs (NPS), which were used for gene expression analysis. Multiple pre-challenge samples were collected from most participants, and the mean of these measurements was used to establish a stable baseline for each participant against which post challenge measurements were compared.

Each study participant was classified in terms of their post-challenge infection and symptom status. Infection status was defined by Bp detection via either ≥1 positive culture or ≥3 positive PCRs at any timepoint from day 6 post-challenge until the first day of azithromycin treatment. Participants were considered symptomatic when exhibiting ≥2 solicited symptoms of which ≥1 respiratory-specific symptom started or worsened with confirmed infection status. Of the 53 participants in our study, 30 (56.6%) had symptomatic infections, 14 (26.4%) had asymptomatic infections, and 9 (17.0%) remained non-infected.

Participants were classified into three distinct clinical outcome groups based on their post-challenge microbiological and symptom status. Infection was defined by positive Bp culture or PCR, while symptomatic infection required the development of respiratory symptoms. Of the 53 participants, 30 (56.6%) developed symptomatic infections, 14 (26.4%) had asymptomatic infections, and 9 (17.0%) remained non-infected (**Figure 1A**).

### Non-infected participants had higher baseline Bp-specific serum IgG titers

Serum antigen-specific IgG titers were measured for the aP antigens PT, FHA, FIM, and PRN. Interestingly, participants who remained non-infected post-challenge had significantly higher pre-challenge titers than infected participants for IgG-PT (all infected *P* = 0.005; asymptomatic false discovery rate [FDR] = 0.079; symptomatic FDR = 0.001) and IgG-FHA (all infected *P* = 0.022; asymptomatic FDR = 0.092; symptomatic FDR = 0.062; **Figure 1C**). Consistent with these higher baseline titers, a larger proportion of non-infected participants had baseline IgG-PT levels above the lower limit of detection (LLOD) compared to both asymptomatic and symptomatic groups. The difference between non-infected and infected participants was not significant for IgG-FIM and IgG-PRN titers, but a trend in the same direction was observed. These findings support an association between higher baseline serum IgG levels against specific Bp antigens and protection from infection.

To validate these baseline protective IgG levels across populations, we independently reanalyzed Bp-specific serum IgG titers from the IMI-2 PERISCOPE study (the first Bp CHIM study)(16, 17). This revealed that non-infected participants in that study also had higher pre-challenge IgG-FHA (*P* = 0.014) and IgG-PRN (*P* = 0.013) titers, while IgG-FIM (*P* = 0.060) and IgG-PT (*P* = 0.310) titers were higher in non-infected participants but did not reach significance (**Figure S1**). Notably, both studies employed an enrollment cutoff for PT-IgG titers; participants were excluded if baseline PT-IgG titers exceeded 20 IU/mL.

### Bp challenge induced a humoral response in symptomatic participants in an antigen-specific fashion

Symptomatic participants exhibited a significant increase in serum IgG against PT, FHA, FIM, and PRN at multiple timepoints post-challenge compared to pre-challenge levels **(Figure 1D**). More specifically, IgG-PT and IgG-FHA titers were significantly higher starting on day 28 and day 21, respectively, and continuing through day 84. IgG-FIM and IgG-PRN titers also rose significantly above baseline on day 21, and IgG-PRN also on day 84. No statistically significant changes were observed in the humoral response for asymptomatic or non-infected participants, indicating that the development of an antigen-specific IgG response was closely associated with a symptomatic infection.

Next, we compared IgG responses after Tdap booster vaccination, based on a reanalysis of our previous study(18), with those observed in symptomatic CHIM participants **(Figure 1E**). This revealed differences in both the timing and magnitude of the IgG responses. Following Tdap booster vaccination, antigen-specific IgG levels peaked rapidly at day 7. In contrast, after Bp challenge, peak titers were delayed, occurring around days 21–28 depending on the antigen, indicating that an infection-induced memory response has slower kinetics. Furthermore, the overall magnitude of the IgG response was lower after Bp challenge than after Tdap booster vaccination, particularly for IgG-FIM.

Within symptomatic participants, we compared pre- to post-challenge antigen-specific IgG titers at all timepoints. Pre-challenge FHA-, PRN-, and FIM-specific IgG titers correlated positively with post-challenge titers for the same antigen, but not for other antigens (**Figure 1F**). This suggests that the boosting of IgG titers is driven by antigen-specific memory rather than a generalized upregulation of humoral immunity.

### Bp challenge decreased systemic mediators independent of challenge outcome

Next, the temporal dynamics of plasma cytokine and chemokine concentrations in response to Bp exposure were examined at each timepoint. Interestingly, all 17 statistically significant changes (FDR<0.05) represented decreases in cytokine or chemokine levels following challenge compared to baseline (**Figure 2A**). This observation held true for all outcome groups (**Figure 2B**). Specifically, the 17 significant changes involved 13 distinct cytokines and chemokines, all of which exhibited decreased concentrations on day 3, with FLT3LG and CCL2 remaining significantly reduced through day 14 (**Figure 2C, S2**). Taken together, Bp exposure caused systemic reductions in cytokine and chemokine concentrations in plasma that were independent of challenge outcome.

**Figure 2.**
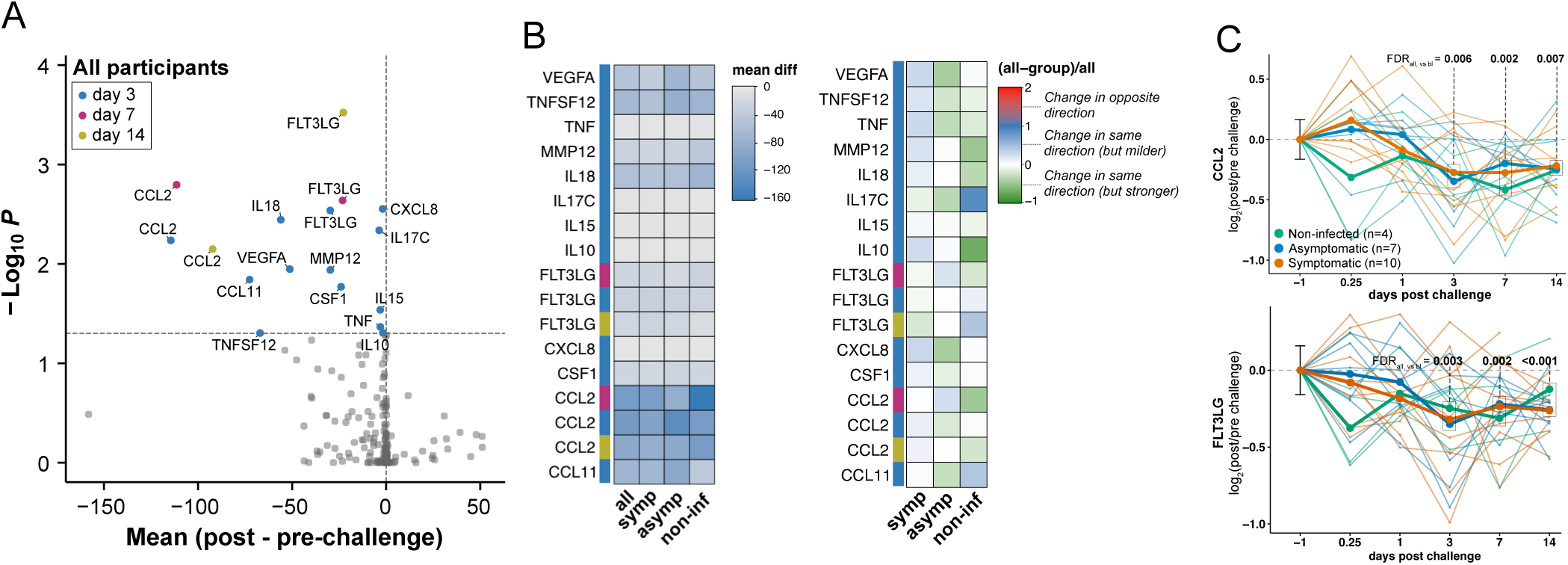
Bp challenge decreased systemic mediators independent of clinical outcome. **A)** Volcano plot showing the plasma cytokine concentration changes of day 0.25, 1, 3, 7, and 14 post/pre-challenge with the mean difference from all participants (x-axis). The −log_10_(FDR) were calculated using a mixed effect model with Dunnett’s multiple comparison test (y-axis). Cytokines that are significantly (FDR < 0.05) decreased are highlighted in blue (day 3), pink (day 7), and yellow (day 14). **B)** Heatmaps depicting the absolute (left) and relative (right) change from baseline for all significantly altered plasma cytokines. Relative values are normalized against the mean of all outcome groups using (all - group) / all. **C)** Plasma CCL2 and FLT3LG concentrations (log_2_ post/pre) over time and coloured per outcome definition. Mean lines are depicted per outcome definition.

### Dynamic changes in blood immune cell frequencies following Bp challenge

The frequencies of 48 distinct blood immune cell subsets were quantified before and after Bp challenge. We identified subsets exhibiting frequency changes in response to Bp challenge, both across all participants and within each outcome group. In total, 24 cell subsets displayed significant changes (FDR < 0.05) at one or more post-challenge timepoints compared to baseline (**Figure 3A**).

**Figure 3.**
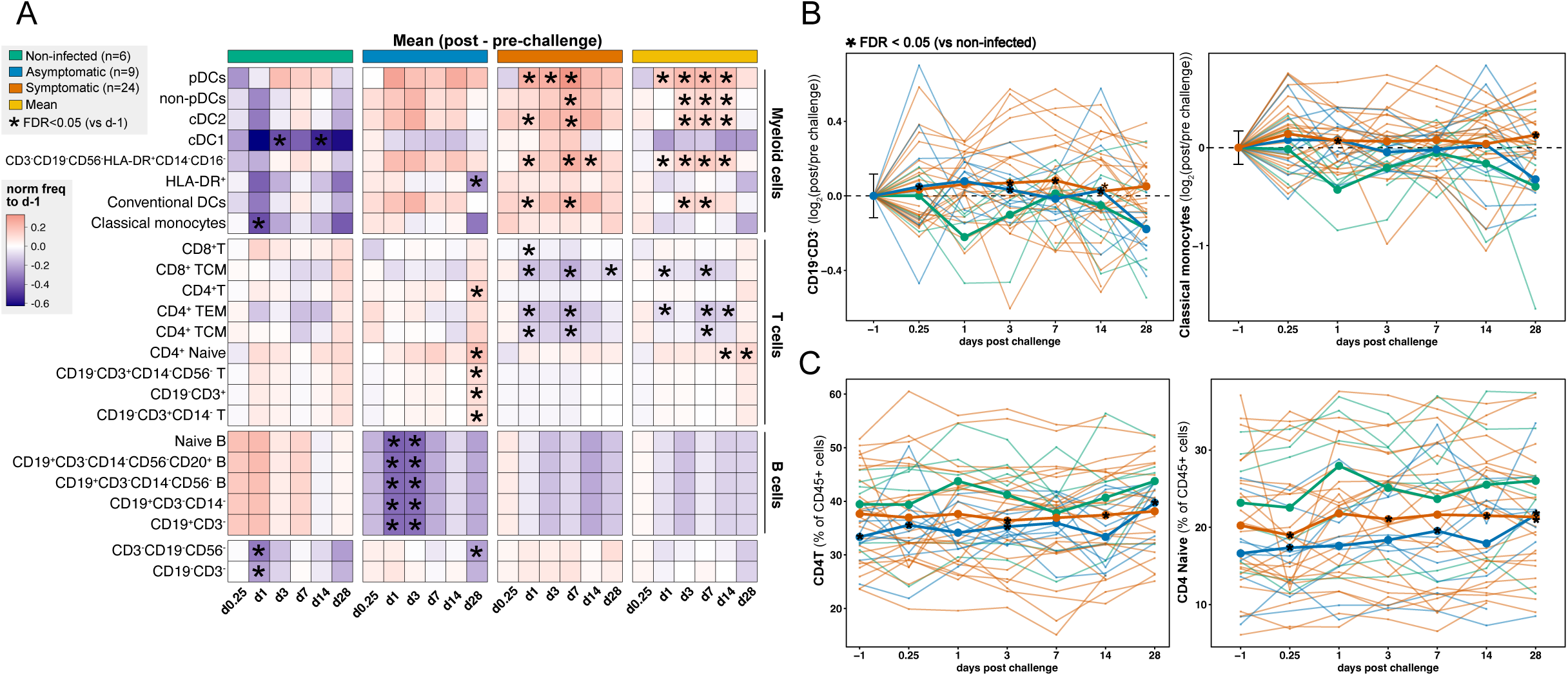
Dynamic and clinically associated changes in blood immune cell frequencies following Bp challenge. **A)** Heatmap showing mean longitudinal changes in PBMC subset frequencies (normalized to pre-challenge levels) for each outcome definition, as well as the overall mean across all outcomes. Only subsets with at least one significant change (FDR < 0.05) are depicted. FDRs were calculated using Dunnett’s multiple comparisons test. **B)** CD19-CD3- (left) and classical monocytes (right) PBMC cell frequencies over time (normalized to pre-challenge frequencies). **C)** CD4T (left) and Naive CD4T (right) PBMC cell frequencies over time. B-C) Mean lines are depicted per outcome definition. Outcome groups were compared at each timepoint using Tukey’s multiple comparisons test and FDR < 0.05 was considered significant (*).

Some changes were outcome-specific. For example, a decrease in the percentage of classical monocytes was observed on day 1 post-challenge only in non-infected participants (FDR = 0.046). In contrast, asymptomatic participants showed increased frequencies of multiple B cell subsets on day 1 and 3 (e.g., naive B cells day 1; FDR = 0.021), while symptomatic participants displayed a reduction in the frequency of central memory CD4L T cells (TCM) on day 1 (FDR = 0.022).

Other changes appeared to be shared among outcome groups, although statistical significance varied. On day 1, symptomatic participants exhibited a significant increase in the frequency of conventional dendritic cells type 2 (cDC2; FDR = 0.040). A similar upward trend was observed in asymptomatic participants (FDR = 0.223). On day 28, asymptomatic participants exhibited elevated frequencies of HLA-DRL cells (FDR = 0.049), reflecting an increase in classical monocytes (FDR = 0.050). A comparable but non-significant trend was observed in non-infected participants for both HLA-DRL cells (FDR = 0.565) and classical monocytes (FDR = 0.586). Additionally, a sustained increase in plasmacytoid dendritic cells (pDCs) on days 3, 7, and 14 post-challenge was observed across all groups, this elevation only reached statistical significance in symptomatic participants.

Outcome-based comparisons further revealed that CD19LCD3L cell frequencies were significantly lower in non-infected participants at multiple timepoints, including day 1 (**Figure 3B, S3A**). This broad population encompasses various myeloid subsets (**Figure S3B**), including monocytes and DCs. Further analysis indicated that the observed differences were primarily driven by reductions in classical monocytes, as this subset was the only myeloid population that showed statistically significant frequency differences across outcome groups (**Figure 3B)** and made up a major fraction of the myeloid cells (**Figure S3B**). In contrast, CD4L T cell frequencies were significantly higher in non-infected participants compared to symptomatic and asymptomatic groups at multiple timepoints, including baseline (**Figure 3C**). Among the more specialized CD4L T cell subsets, naïve CD4L T cells exhibited a similar outcome-associated pattern. To conclude, Bp challenge dynamically changed blood immune cell frequencies in a challenge outcome-dependent manner.

### Limited transcriptional changes in PBMCs following Bp challenge

RNA-Seq of bulk PBMCs was performed to assess peripheral differences following Bp challenge. Strikingly, time-course analysis of symptomatic participants revealed no differentially expressed genes (DEGs) on days 0.25 (4-6 hours), 1, 3, and 7 post-challenge relative to baseline (**Figure S4**). In contrast, by day 14, 34 DEGs were identified (31 downregulated and 3 upregulated; **Figure 4A**), many of which were B cell-associated transcripts such as *CD24, CD79B, SPIB,* and *IGHM* (**Figure 4A-B**). To investigate if this transcriptomic signal reflected changes in cellular composition, we analyzed circulating B cell frequencies and found a reduction in the percentage of B cells at day 14 compared to baseline in symptomatic participants (*P*=0.085; FDR=0.273; **Figure 4C**). Further B cell subset analysis demonstrated decreases across all circulating B cell subsets, with the most pronounced reductions in naive B cells (*P*=0.092; FDR=0.290) and memory (*P*=0.137; FDR=0.307), and the least in proliferating B cells (*P*=0.333; FDR=0.974). Within proliferating and memory B cell subsets, stratification into activated versus antibody-secreting phenotypes revealed that activated subsets were reduced at day 14 (activated memory: *P*=0.105; FDR=0.293; activated proliferating: *P*=0.043; FDR=0.608), whereas antibody-secreting B cells were increased (antibody-secreting proliferating: *P*=0.168; FDR=0.411; antibody-secreting memory: *P*=0.085; FDR=0.961). Although these cellular changes did not reach statistical significance, the directional trend aligns with the observed reduction in B cell-specific transcripts, with downstream subset analysis suggesting this overall decrease is primarily driven by naive, activated memory, and proliferating B cell subsets. By day 28, these transcriptional signatures had returned to baseline levels (**Figure S4**). Together, these findings indicate that Bp challenge induced minimal transcriptional changes in PBMCs overall.

**Figure 4.**
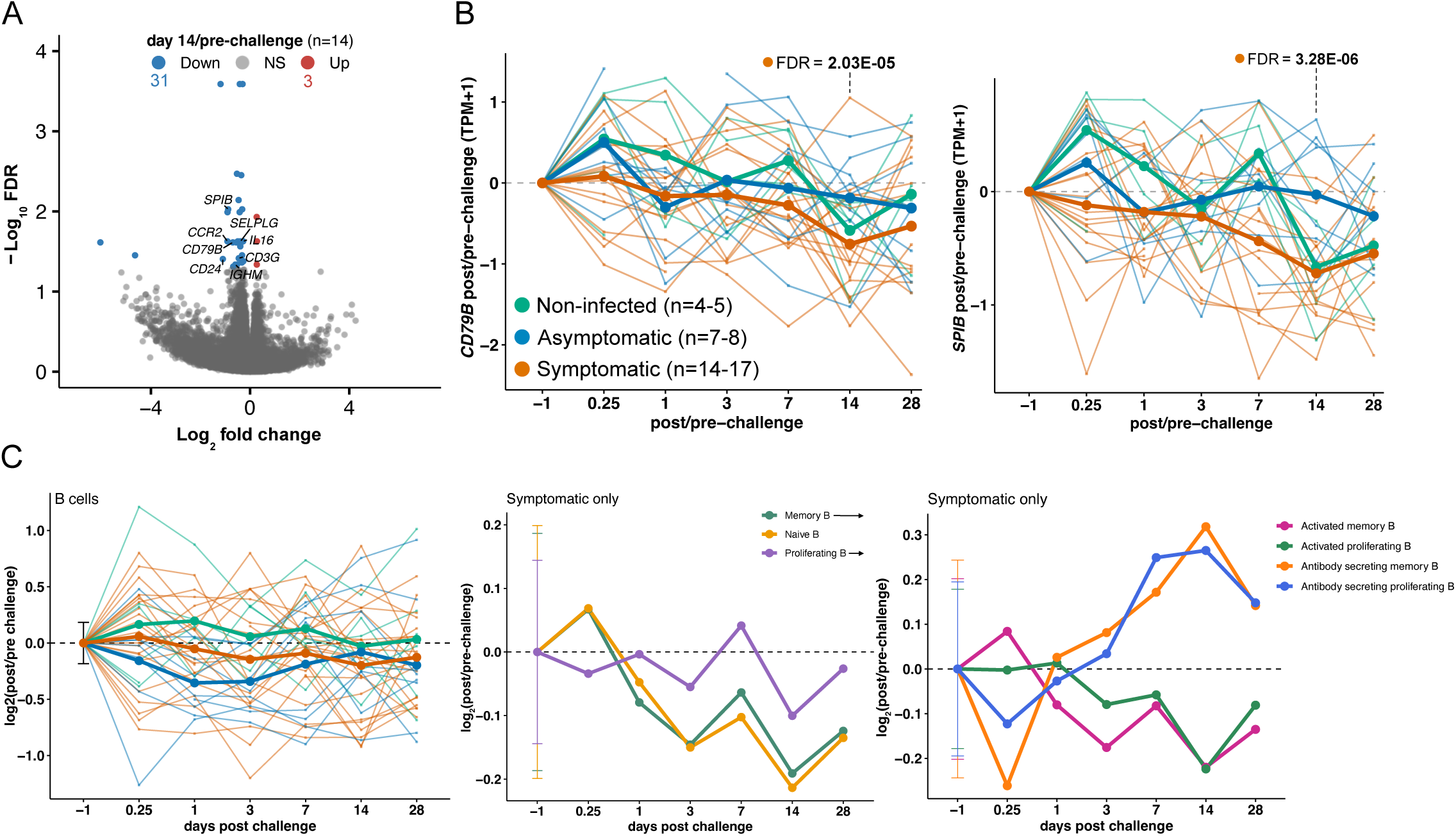
Limited transcriptional changes in PBMCs following Bp challenge. **A)** Volcano plot showing the log_2_(fold change) and −log_10_(FDR) of the EdgeR differentially expressed genes (DEG) in PBMCs of symptomatic people on day 14 vs. pre-challenge with downregulated genes in blue and upregulated genes in red (FDR < 0.05). **B)** Pre-challenge normalized CD79B (left) and SPIB (right) gene expression (TPM+1) over time in PBMCs. **C)** Frequencies of B cells (left) and B cell subsets (middle: memory, naïve, and proliferating; right: activated and antibody-secreting, memory and proliferating) over time, normalized to pre-challenge frequencies. In the left panel, mean lines are shown for each outcome definition. In the middle and right panels, only symptomatic participants are shown, and mean lines represent each B cell subset. P values were calculated by multiple two-tailed Wilcoxon matched-pairs tests. FDRs were calculated using Dunnett’s multiple comparisons test.

### Antigen-specific T cell activation and polarization in the blood were largely unaffected by Bp exposure

To determine antigen-specific T cell immunity before and after Bp challenge, T cell activation and polarization assays were performed. PBMCs were stimulated with peptide pools containing aP or non-aP Bp antigens. T cell activation was measured using an activation-induced marker (AIM) assay(19), which determined the percentage of CD4L T cells that co-expressed the T cell activation surface markers TNF receptor superfamily member 4 (OX40) and interleukin 2 receptor alpha (CD25).

Already at baseline, both DTaP-primed and DTwP-primed participants showed aP-specific and non-aP-specific T cell responses (**Figure 5A**). This suggests that aP-primed participants were likely exposed to non-aP Bp antigens at some point, likely through environmental exposure to Bp with subclinical infection–a phenomenon previously described by da Silva *et al.*(20). Remarkably, no clinical outcome group showed a significant boost in T cell immunity against aP or non-aP Bp antigens post-challenge at any of the timepoints measured (**Figure 5B** for TT corrected and **Figure S5A** for DMSO corrected data). Furthermore, no differences in T cell activation were observed between the different outcome groups. As previous research has shown that aP-specific T cell activation is increased upon intramuscular Tdap booster vaccination(18, 21), we conclude that nasal Bp exposure had less of an effect on blood T cell immunity against Bp.

**Figure 5.**
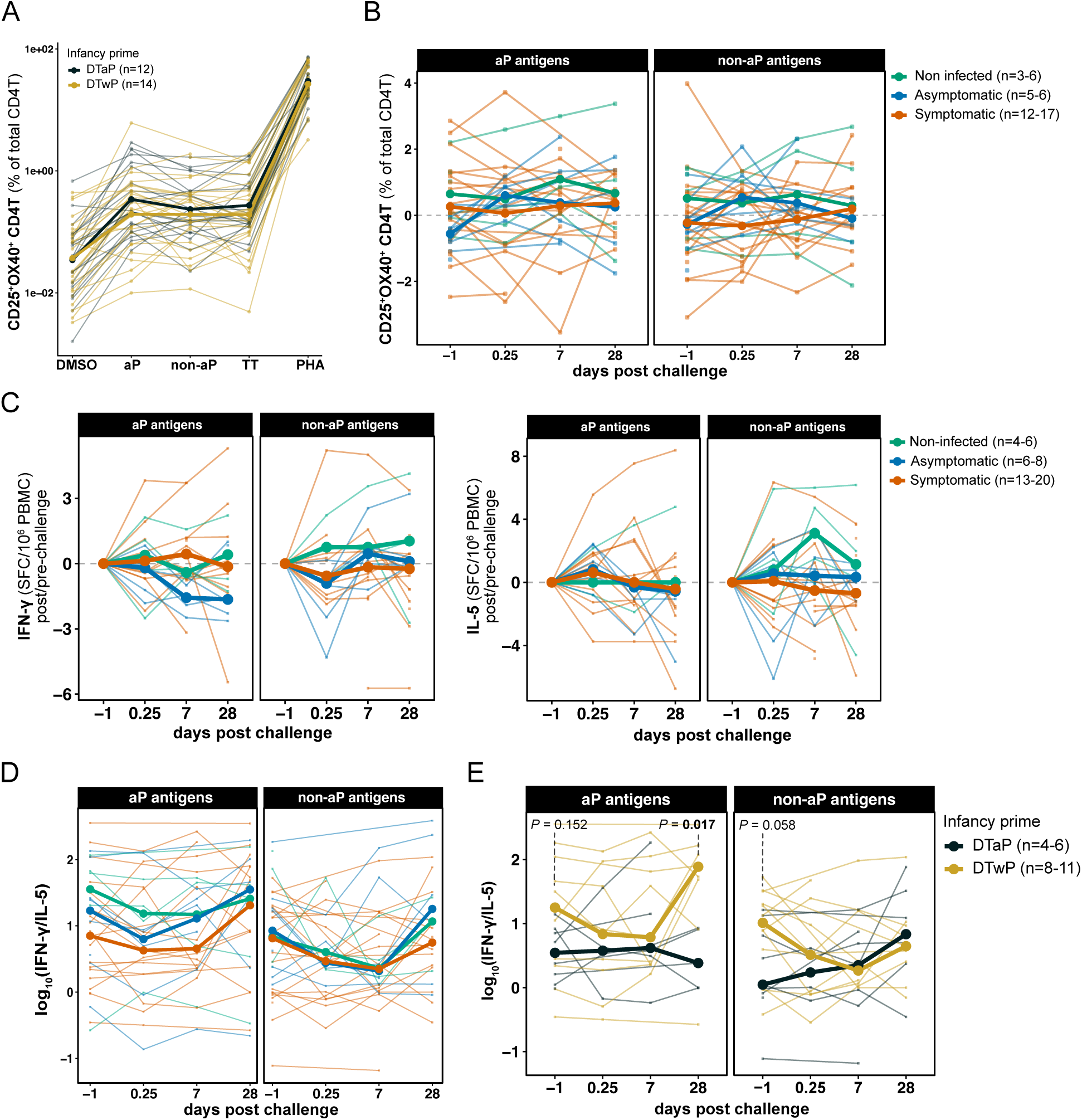
Antigen-specific T cell activation and polarization in the blood were unaffected by Bp exposure. **A)** Activation-induced marker (AIM) assays: PBMCs were stimulated with 1 μg/mL pertussis antigen megapools (containing aP or non-aP Bp vaccine antigens), tetanus peptide megapools, DMSO (negative control), or PHA (positive control) for 18–24 h. CD25LOX40L CD4L T cells were quantified by flow cytometry and expressed as a percentage of total CD4L T cells. Mean lines are shown per primary vaccine group. **B)** TT-normalized frequencies of CD25LOX40L CD4L T cells. Mean lines are shown per outcome group. P values were calculated using multiple two-tailed Wilcoxon matched-pairs tests. **C)** IFN-γ and IL-5 producing cells (spot-forming cells, SFC) were measured by Fluorospot following 14-day stimulation with aP or non-aP Bp megapools. Data are shown normalized to pre-challenge measurements. **D)** The IFN-γ/IL-5 ratio was used as a measure of Th1/Th2 polarization and is shown over time and coloured by clinical outcome. **E)** IFN-γ/IL-5 ratios stratified by primary pertussis vaccine group in symptomatic participants. P-values were calculated by multiple two-tailed Mann-Whitney tests. Median lines are shown in panels C-E.

T cell polarization was evaluated by quantifying cytokine-secreting cells from blood following *in vitro* stimulation with aP or non-aP Bp vaccine antigens with a 14-day expansion period. IFN-γ/IL-5 was used as a marker for Th1/Th2 polarization, respectively(22). Longitudinal analysis revealed that Bp challenge did not significantly increase the number of IFN-γ or IL-5-producing cells in response to either aP or non-aP Bp antigen stimulation in any of the challenge outcome groups (**Figure 5C-D, S5C**). Although no significant differences were observed in Th1/Th2 polarization ratio (**Figure 5D**), symptomatic participants exhibited more aP-specific IFN-γ-producing T cells at baseline compared to non-infected participants (FDR=0.041), and at days 0.25 (FDR=0.043) and 7 (FDR=0.001) post-challenge compared to asymptomatic participants (**Figure S5C**).

Consistent with previous Tdap booster vaccination studies using aP-peptide stimulations(18, 20–24), the Th1/Th2 polarization ratio in response to aP antigens in symptomatic participants trended higher in wP-primed compared to aP-primed participants at baseline (P=0.152) and was significantly higher 28 days post-challenge (P=0.016; **Figure 5E**). After stimulation with non-aP Bp peptides, a trend in the same direction was observed at baseline (P=0.058). These differences within symptomatic participants were driven by both elevated IFN-γ and reduced IL-5 production in wP-primed participants, with significantly more IFN-γ-producing T cells at baseline after aP stimulation (P=0.009; **Figure S5D**). Collectively, these findings suggest that Bp challenge does not significantly alter blood T cell polarization.

### Bp challenge induced a nasal inflammatory response on day 7 in participants who subsequently developed symptoms

To characterize the mucosal immune response to nasal Bp exposure, bulk RNA sequencing was performed on NPS collected at baseline and on days 1, 3, 7, 14, and 28 post-challenge. Differential expression analysis was conducted within each challenge outcome group to identify genes with altered expression following challenge relative to baseline. The most pronounced transcriptional changes were observed on day 7 in symptomatic participants (**Figure 6A**), comprising 1,196 DEGs (FDR<0.05), of which 1,150 were upregulated and 46 were downregulated. The upregulated DEGs on day 7 accounted for 78% of all DEGs identified across outcome groups and 92% of those within the symptomatic group. Notably, no upregulated DEGs were detected on day 7 in asymptomatic or non-infected participants.

**Figure 6.**
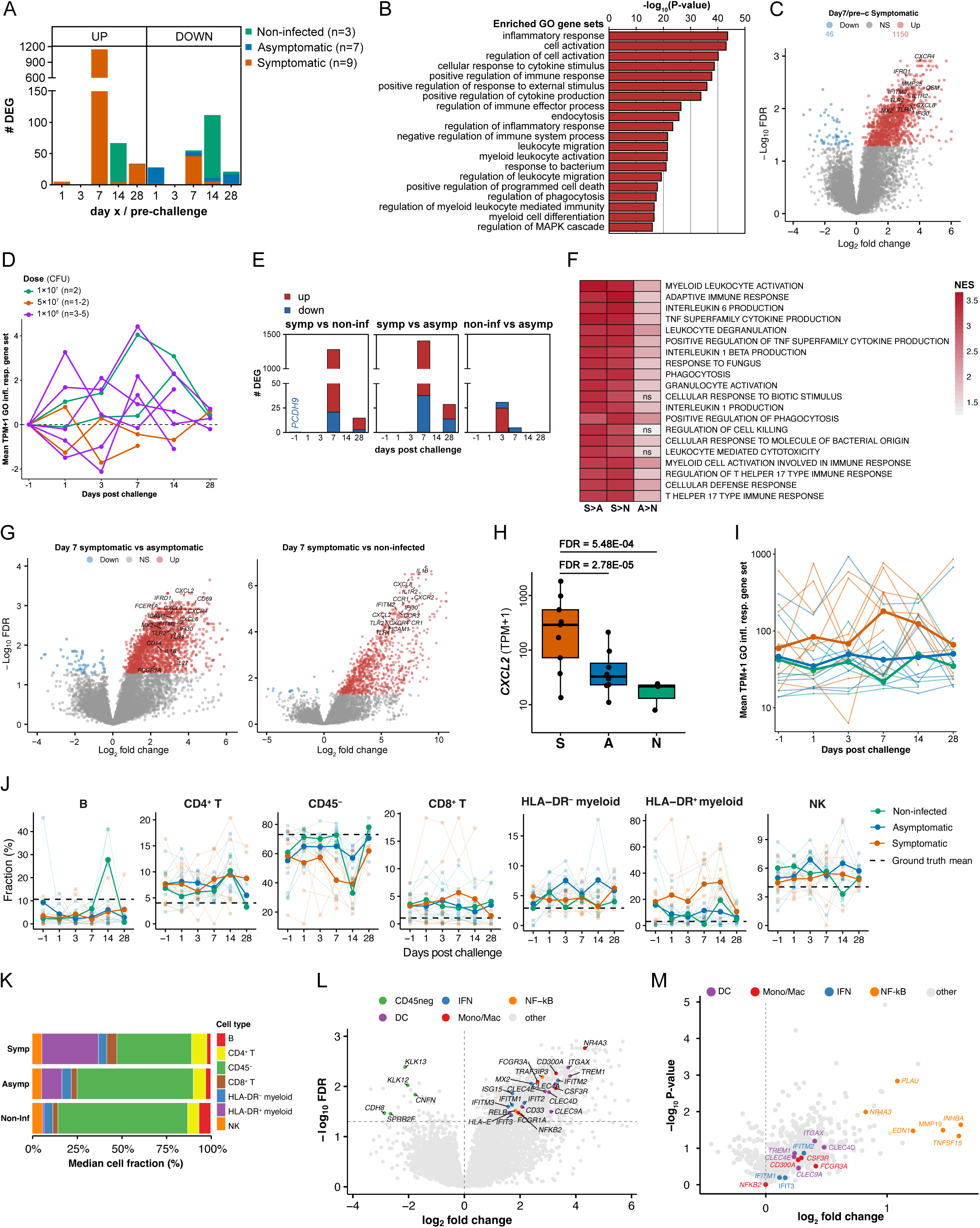
Bp challenge induced a nasal inflammatory response on day 7 in participants who subsequently developed symptoms. **A)** The number of DESeq2 differentially expressed genes (DEG; FDR < 0.05) in NPS of the longitudinal analysis per timepoint and outcome group. **B)** The most significantly enriched gene sets identified by Gene Ontology (GO) enrichment analysis using the 1,150 genes that were upregulated in NPS of symptomatic people on day 7 vs. pre-challenge. **C)** Volcano plot showing the log_2_(fold change) and −log_10_(FDR) of the DEG in NPS of symptomatic people on day 7 vs. pre-challenge with downregulated genes in blue and upregulated genes in red. **D)** Pre-challenge normalized CXCR4 gene expression (TPM+1) over time in NPS of symptomatic people. **E)** The number of DEG (FDR < 0.05) in NPS of each outcome comparison per timepoint. **F)** Heatmap of normalized enrichment scores (NES) for the top 20 (highest absolute NES) GO biological process gene sets in NPS across each outcome comparison on day 7. Cell colors range from light grey to dark red, indicating increasing positive NES values. Gene sets enriched with FDR > 0.05 are marked “ns”. **G)** Volcano plots showing the log_2_(fold change) and −log_10_(FDR) of the DEG in NPS of symptomatic vs asymptomatic people (left) and symptomatic vs non-infected people (right) on day 7 with downregulated genes in blue and upregulated genes in red. **H)** *CXCL2* gene expression (TPM+1) in NPS on day 7 per outcome definition. The upper and lower hinges correspond to the first and third quartiles and the middle line represents the median. **I)** Expression (mean TPM+1) of genes significantly upregulated on day 7 in symptomatic participants overlapping with the inflammatory response (GO:0006954) gene set. **J)** Predicted NPS cell frequencies using CIBERSORTx RNA deconvolution. Mean per outcome group is shown. **K)** Mean predicted cell type composition of NPS at day 7 by outcome group. **L)** Volcano plot highlighting cell-type and immune response-related markers among the NPS DEG of symptomatic people on day 7 vs baseline. **M)** Volcano plot showing the log_2_(fold change) and −log_10_(P-value) of the DEG in HLA-DR+ myeloid cells of symptomatic people on day 7 vs. pre-challenge.

Next, the upregulated DEGs on day 7 in symptomatic participants were analyzed for the functional properties underlying their coordinated expression (**Figure 6B**). Gene set enrichment analysis (GSEA) based on Gene Ontology (GO) biological process annotations revealed a predominance of immunologically related gene sets. The five most significantly enriched gene sets were inflammatory response (GO:0006954), cell activation (GO:0001775), regulation of cell activation (GO:0050865), cellular response to cytokine stimulus (GO:0071345), and positive regulation of immune response (GO:0050778). The most significantly upregulated DEGs included members of these immune-related gene sets, such as *CXCR4, CXCL8, TLR2, TLR4, MMP25, MX2, IFRD1, IFITM2, IL1R2,* and *OSM* (**Figure 6C, S6A**), and their induction appeared independent of bacterial dose (**Figure 6D)**.

To determine whether this transcriptional response was specific to symptomatic individuals or if immune-related genes were similarly upregulated in other outcome groups without reaching statistical significance, we performed outcome-specific comparisons at each timepoint (**Figure 6E**). At baseline, *PCDH9* was the only DEG and was expressed higher in non-infected individuals compared to those who became symptomatic. The absence of immune-related DEGs at baseline suggests that pre-challenge nasal gene expression is not a major determinant of later differences. Following Bp challenge, the majority of DEGs were identified on day 7, with a substantial number of upregulated genes in symptomatic individuals compared to asymptomatic (1,374 genes) or non-infected participants (1,264 genes). In contrast, only five DEGs (*FCN1, OSM, FFAR2, IL1B,* and *PROK2*) were observed between the asymptomatic and non-infected groups (**Figure S6B**). A relatively small number of genes were downregulated in symptomatic individuals compared to asymptomatic individuals (38 genes) or non-infected individuals (21 genes). Gene set enrichment analysis (GSEA) was conducted to test whether similar immune-related gene sets were enriched in symptomatic versus asymptomatic or non-infected individuals (**Figure 6F**). Indeed, GSEA showed that symptomatic participants exhibited strong significant enrichment (FDR < 0.05, high normalized enrichment scores (NES)) for the same immune-related gene sets, such as phagocytosis and adaptive immune response, whether they were compared to asymptomatic or non-infected participants. Furthermore, when comparing the asymptomatic group to the non-infected group, these same gene sets displayed lower yet also positive NES, but higher P-values. Genes with immune-related functions that were found to be upregulated in symptomatic participants versus asymptomatic or non-infected participants included *IL1B, CXCL8, IL1R2, CXCR2, CXCL2, CCR1, ICAM1, and CR1* (**Figure 6G-H**).

Interestingly, the GSEA results and expression profiles indicated a graded activation pattern across challenge outcome groups: many immune-related gene sets were enriched in symptomatic individuals over asymptomatic and non-infected participants, and the same sets were modestly enriched in asymptomatic over non-infected individuals. Moreover, the five DEGs distinguishing asymptomatic from non-infected participants displayed progressively increasing expression from non-infected to asymptomatic to symptomatic individuals (**Figure S6B**). Consistently, the mean expression of genes significantly upregulated on day 7 in symptomatic participants that overlapped with the inflammatory response gene set (GO:0006954; 104 genes) followed the same gradient across outcome groups (**Figure 6I**). In conclusion, Bp challenge elicits a robust nasal transcriptional immune response on day 7, which is strongest in symptomatic individuals but detectable as a gradient across challenge outcomes, suggesting a continuum of mucosal inflammatory activation associated with symptom development.

### Increased nasal HLA-DR^+^ myeloid cell infiltration and cell-type-specific inflammatory responses on day 7 in symptomatic individuals

Next, to investigate the cellular origins of the transcriptional upregulation of immune-related genes in NPS samples from symptomatic participants on day 7, we performed bulk RNA-Seq deconvolution. While algorithms to deconvolute RNA-Seq data into cell frequencies from blood samples are frequently used and have been shown to be reliable, there is much less data available from nasal samples. To determine which method to use and how trustworthy it is, we first benchmarked different transcriptomic deconvolution approaches against a newly generated reference dataset consisting of flow cytometry and RNA-Seq data from NPS samples (unchallenged healthy people; n = 13; **Figure S6C-E**). We found that CIBERSORTx(25) used with a nasal immune cell signature matrix derived from Ramirez et al.(26) performed best (**Figure S6F-H**); therefore, this approach was used in all subsequent analyses.

Applying this method to estimate immune cell compositions in Bp CHIM NPS samples revealed an increased proportion of HLA-DR^+^ myeloid cells (including DCs, monocytes, and macrophages) from 18.2% at baseline to 36.0% on day 7 in symptomatic participants (**Figure 6J**). Additionally, a higher proportion of HLA-DR+ myeloid cells was observed in symptomatic individuals compared with those from asymptomatic and non-infected individuals (36.0% vs 6.0% and 1.0%, respectively; **Figure 6K**).

The expansion of HLA-DR^+^ myeloid cells was accompanied by a relative decrease in CD45L cell types (**Figure 6J**), notably erythroblasts and epithelial populations including ciliated, basal, and secretory cells. Consistent with this shift, the genes upregulated on day 7 in symptomatic participants included classical DC markers (e.g., *CLEC4A, CLEC4D, CLEC4E, CLEC9A, TREM1, HLA-E, CD33,* and *ITGAX)* as well as monocyte and macrophage markers (e.g., *FCGR1A, FCGR3A, LILRA, LILRB, NR4A3, CSF3R, CD300A,* and *CD300B;* **Figure 6L**). Moreover, the downregulated genes on day 7 contained multiple epithelial-associated genes (e.g., *KLK13, KLK12, CNFN, SPRR2F,* and *CDH8*).

While many of the upregulated DEGs likely reflect changes in cell composition, several are also transcriptionally induced by inflammatory cues, such as *NFKB2* and *RELB* (indicative of NF-κB signaling), and interferon-stimulated genes (*IFITM1, IFITM2, IFITM3, IFIT2, IFIT3, ISG15, MX2*), which could be either activated across a specific or multiple immune cell subsets. To distinguish between changes induced by differences in cell frequencies versus changes in activation status, we deciphered the changes in gene expression per cell type at day 7 relative to baseline using CIBERSORTx’s gene expression profiles (GEP) function. Differential expression analysis of the HLA-DR^+^ myeloid population’s GEPs in symptomatic participants revealed that NF-κB signaling genes such as *MMP19*(27), *TNFSF15*(28), *PLAU*(29), *NR4A3*(30), *INHBA*(31), and *EDN1*(32) were upregulated within HLA-DR^+^ myeloid cells on day 7 relative to baseline (**Figure 6M**, *P*<0.05). Within the HLA-DR^+^ myeloid population, no clear changes were observed in IFN-associated genes nor in myeloid subset markers. Taken together, these data suggest that the increase in inflammatory gene expression on day 7 in symptomatic participants is likely driven by an increase in DCs, monocytes, and macrophages in the nasal mucosa, combined with a specific NF-κB-mediated activation of these HLA-DR^+^ myeloid cells.

## Discussion

Our study provides an integrated view of systemic and mucosal immune responses to Bp challenge, focusing on changes in cellular responses in terms of composition, gene expression, and activation profiles. By combining longitudinal sampling from both blood and nasal compartments, we identified clinical outcome-specific immune responses. Our analyses reveal that (i) non-infected participants exhibited higher baseline Bp-specific serum IgG titers; (ii) serum IgG responses were particularly induced in symptomatic individuals in an antigen-specific fashion; (iii) infection-induced serum IgG responses displayed slower kinetics and a lower overall magnitude compared to the rapid day 7 peak observed following Tdap booster vaccination; (iv) participants who developed symptoms showed nasal inflammation with an increase in HLA-DR^+^ myeloid cells and cell-type-specific NF-κB activation on day 7 post-challenge; (v) blood immune cell frequencies shifted dynamically, with some changes linked to challenge outcomes; (vi) plasma cytokine concentrations decreased independent of challenge outcome; (vii) transcriptional changes occurred predominantly in the nasal mucosa rather than in the blood; and (viii) antigen-specific T cell activation and polarization in the blood were largely unaffected by Bp exposure. Together, these findings provide novel insights into correlates of protection and symptom development following human pertussis challenge.

Our finding that non-infected participants had significantly higher pre-challenge serum IgG titers against PT and FHA in this study and against FHA and PRN in the IMI-2 PERISCOPE study(16, 17) suggests that circulating antigen-specific antibodies play a key role in preventing infection. These results align with other clinical studies: Storsaeter *et al.*(33) showed that higher PT-IgG, FIM-IgG, and PRN-IgG titers correlate with clinical protection, Deen *et al.*(34) demonstrated that PRN-IgG and FHA-IgG ELISA values above a certain threshold prevented clinical pertussis. Similarly, de Graaf *et al.* found that uncolonized volunteers had higher pre-challenge serum IgG concentrations against PT, PRN, and FHA compared to those who became colonized(35). Cherry *et al.* also found that PRN-IgG and PT-IgG titers were significant in providing protection(36). Notably, both the North American and PERISCOPE Bp CHIM studies employed an enrollment cutoff, excluding participants with baseline PT-IgG titers >20 IU/mL, respectively. While this ensured a susceptible study cohort, removing this criterion might have enabled more detailed insights into (absolute) protective thresholds. We emphasize that in the general population (with a broader range of baseline titers), the protective effect of these antibodies would likely be even more pronounced.

Interestingly, the correlation between baseline and post-challenge IgG titers in infected participants suggests that memory B cell pools are engaged in an antigen-restricted manner. The fact that IgG titers did not increase in non-infected participants post-challenge suggests that pre-existing IgGs induce local pathogen elimination without promoting a systemic response. Therefore, serological profiles could be used to predict protection against infection. Since protection is known to be mediated not only by humoral immunity but also by cellular immunity, modeling based solely on serology profiles will likely not achieve a 100% success rate.

In this study, seroconversion was observed in infected participants (symptomatic and asymptomatic)(15), confirming that a systemic humoral response can be induced in the absence of symptoms(16, 37). However, we found that the development of symptoms and antigen-specific IgG responses were interrelated; symptomatic participants were nearly twice as likely to seroconvert (21/30, 70.0%) as those who remained asymptomatic (5/14, 35.7%)(15). Furthermore, the significantly higher magnitude of the humoral response in symptomatic participants likely reflects increased antigenic stimulation, as these participants exhibited a higher bacterial load compared to other outcome groups(15), a correlation supported by previous findings(13, 14).

Asymptomatic infections have been documented in vaccinated individuals, indicating that silent infection and potential transmission can occur(16, 38–40). The presence of antigen-specific T cell responses against non-aP Bp antigens in aP-primed individuals further supports the existence of subclinical exposure(20). Therefore, future vaccines must also prevent transmission from asymptomatic carriers to achieve herd protection. Such a strategy has been successful for respiratory pathobionts, such as *Streptococcus pneumoniae, Neisseria meningitidis*, and *Haemophilus influenzae,* in which vaccines interrupt the carrier state to prevent disease spread(41, 42). Interestingly, harmless infection can also boost natural immunity, as some participants seroconverted, and protect against severe disease(16). Further research is required to directly compare the quality and durability of immune memory from asymptomatic versus symptomatic Bp infections in humans.

We observed differences in the timing and magnitude of the humoral response between participants challenged with Bp and those who received a Tdap booster vaccination. Following Tdap booster, antigen-specific IgG titers peaked rapidly on day 7. In contrast, after Bp inoculation, peak titers were delayed, occurring around days 21–28 depending on the antigen, reflecting that a more natural memory response has slower kinetics. Furthermore, the overall magnitude of the IgG response was lower after Bp inoculation than after Tdap booster. This pattern aligns with previous findings from one of the PERISCOPE Bp CHIM studies(16, 17). Possible explanations for these differences include: (i) lower systemic antigen exposure after Bp inoculation compared to intramuscular Tdap vaccination, potentially due to local immune regulation at the mucosal site limiting systemic IgG amplification, and (ii) differences in antigen presentation and immune stimulation between live bacterial exposure and inactivated vaccine formulations.

Surprisingly, systemic immune responses detectable in blood following Bp challenge were modest. Plasma cytokine and chemokine concentrations decreased after challenge regardless of outcome, and PBMC transcriptional changes were limited, with transient reductions in B cell-associated transcripts observed only on day 14 in symptomatic participants. Furthermore, antigen-specific T cell activation and polarization were not altered following challenge, despite robust expansion after intramuscular Tdap booster vaccination in wP-primed participants in prior studies(18, 21). In contrast to the blood compartment, mucosal immune responses were strong in symptomatic participants.

This disparity suggests that Bp elicits a predominantly mucosal response. The lack of systemic T cell engagement may occur because antigen-specific memory T cells reside within the nasal mucosa and are therefore not captured in the blood. In support of this, de Graaf *et al.* reported no significant changes in the percentage of Th1 (IFN-γ), Th2 (IL-4, 5, 13), Th17 (IL-17A/F), or Th22 (IL-22) CD4^+^ T cells in the peripheral blood of colonized volunteers at day 28 post-challenge(35). This hypothesis is further supported by mouse studies demonstrating that CD4^+^ lung tissue-resident memory T cells (T_RM_) mediate adaptive immunity induced by previous infection with Bp. These Bp-specific Th1/Th17 cells accumulated in the lungs during initial infection and significantly expanded through local proliferation following reinfection(43). In addition, McCarthy *et al.* showed that CD4^+^ T_RM_ cells are recruited and persist in the human respiratory tissues after primary pertussis vaccination, while CD4^+^ T cell responses in blood were weak or undetectable(44).

The absence of a broader systemic immune response may reflect both immune evasion strategies of Bp and compartmentalization of the host response. Although virulence factors such as PT and ACT can dampen systemic immunity(45, 46), an alternative explanation is that infection remains largely confined to mucosal and draining lymphoid compartments, limiting systemic immune activation. In contrast, systemic immune activation following Tdap vaccination may reflect the absence of PT-mediated immune modulation and its intramuscular route of administration.

Additionally, Jazayeri *et al*. demonstrated that nasal immunization with ciprofloxacin-treated Bp, which retains toxin structure and activity, did not induce systemic inflammatory responses in mice while still conferring protection against infection(47). Given that systemic immune responses have been associated with adverse events in mice(48), a vaccine that prevents infection without eliciting systemic responses may be safer.

In contrast to the blood compartment, mucosal transcriptional immune responses were robust in symptomatic participants, indicating that symptom development is primarily linked to local rather than systemic immune activation. Nasal gene expression signatures showed strong activation of innate immune responses, including IFN and NF-κB signaling, accompanied by a predicted increase in monocytes, macrophages, and DCs. This expansion likely reflects both recruitment of circulating monocytes that differentiate locally and proliferation of resident macrophages and DCs. Consistent with this, mouse studies have reported a peak in lung macrophages(49) and infiltrating CD11c^+^CD8α^+^ DCs(50) following aerosol Bp challenge and showed that these subsets play an important role in immunity to Bp and bacterial clearance(9). While these responses support protective immunity, they also promote tissue inflammation and pathology, thereby contributing to symptom development. Reduced epithelial-associated transcripts further support disruption of mucosal homeostasis during symptomatic infection. Finally, the increase in HLA-DR^+^ myeloid cells observed in the nasal mucosa paralleled changes in blood, suggesting that infection-induced myelopoiesis in the bone marrow may underlie the systemic and local increase in myeloid cells.

This study has several limitations. First, group sizes were limited, which particularly affected comparisons involving non-infected participants and restricted analyses of priming status across outcome groups. Second, the challenge strain expressed PRN, whereas many circulating strains are PRN-deficient, and immune responses may differ accordingly. Finally, while bulk transcriptional analyses on the nasal mucosa followed by computational deconvolution revealed differences in mucosal immunity, single-cell and spatial analyses would provide a higher resolution picture of cell-type–specific activation states and interactions.

In summary, this study demonstrates that protection against Bp infection is associated with pre-existing serum IgG titers, while symptom development is linked to localized nasal inflammation and likely myeloid expansion. Systemic immune responses to Bp exposure were limited and in contrast to responses induced by Tdap booster vaccination. These findings highlight the importance of mucosal immunity in pertussis and provide correlates of protection to guide the rational design and evaluation of next-generation vaccines aimed at preventing both disease and transmission.

## Methods

### Trial design and human participants

Healthy adults (n=53), aged 18-40 years, who were primed during infancy with either DTwP or DTaP vaccines were recruited for the study (**Figure 1A, Table S1-3**). Participants were excluded when they received a Bp booster vaccination within the previous five years, had a reported history of laboratory-confirmed Bp infection, or had serum PT-IgG titers >20 IU/mL. Full inclusion and exclusion criteria are listed at(15). All participants provided written informed consent prior to enrollment and study procedures. This study was conducted in accordance with the Declaration of Helsinki and the International Conference on Harmonisation guidelines for Good Clinical Practice (ICH GCP), and the protocol was approved by the IWK Health’s Research Ethics Board (REB# 1026718). On day 0, participants were intranasally challenged with 0.2 mL Bp (US Bp D420 strain; PRN-producing) with one of the four dose groups 5×10L (n=10), 1×10L (n=22), 5×10L (n=12), or 1×10L (n=9). All participants were carefully monitored in an inpatient challenge unit at the Canadian Center for Vaccinology (CCfV) in Halifax, Nova Scotia. Asymptomatic and non-infected participants were discharged on day 16 and completed a 5-day course of azithromycin as outpatients. Symptomatic participants received a 5-day course of azithromycin initiated 24-48 h after symptom onset and were discharged following treatment completion. All participants were subsequently followed up through scheduled clinic visits and phone consultations. Longitudinal blood and nasal samples were collected before and after Bp-challenge (Figure 1B). Study outcomes were categorized as symptomatic infection, asymptomatic infection, or non-infected based on diagnostic testing and symptom manifestation. Infection status was defined by Bp detection via either ≥1 positive culture or ≥3 positive PCRs between day 6 post-challenge and the first day of azithromycin treatment(15). Participants were categorized as symptomatic when exhibiting ≥2 solicited symptoms of which ≥1 respiratory-specific symptom started or worsened with confirmed infection status.

### Serology

Bp serum antibodies were quantified using a validated enzyme-linked immunosorbent assay (ELISA) with reference sera standardized against U.S. Reference Pertussis Antiserum (Human) Lot 3.20. Bp antigens were adsorbed onto polystyrene flat-bottom plates (Nunc Polysorp) at the following concentrations: 0.5 µg/mL PT (List Biological Laboratories Inc), 0.5 µg/mL FHA (Connaught), 1.5 µg/mL PRN (Sanofi), and 0.75µg/mL FIM (Sanofi), and incubated for 16-24 h. All incubations, including the coating step, were performed on a rotator/incubator at 28°C and 100 RPM. Between steps, plates were washed three times with PBS containing 0.005% PPG, 0.5% BSA, and 0.05% Tween-20 (Sigma). Reference, control, and test sera were serially diluted in incubation buffer. Then, 100 µL of each dilution was added to the antigen-coated wells and incubated for 2.5 h. Subsequently, 100 µL of phosphatase-labeled goat anti-human IgG (KPL) was added to all wells and incubated for 16-24 h. Finally, 100 µL of 1 mg/mL p-nitrophenyl phosphate (Sigma) in substrate buffer (1 M Tris/0.3 mM MgCl2, pH 9.8) was added, and after 1 h, absorbance was measured at 405 nm using a Biotek Cytation 1 Imaging Reader. Total serum IgG concentrations against PT, FHA, PRN and FIM were calculated reference line software (Gen5, v3.11) and expressed as international units per mL (IU/mL) for all antigens except FIM which was in ELISA units per mL (EU/mL); the lower limits of quantification (LLQ), were 10 IU, 10 IU, 10 IU, and 16 EU, respectively.

To reduce the impact of tied values on rank-based correlations, we repeatedly added small random jitter (0–0.001) to tied observations in the baseline data and recalculated Spearman correlations (n = 1000 iterations). The average correlation coefficient and p-value across all iterations were reported. This Monte Carlo jittering approach provides a stable estimate of the association while minimizing the effect of ties.

### Flow cytometry PBMC cell frequencies

One million PBMC were stained with 1% Zombie UV fixable viability dye (BioLegend) in PBS and incubated for 15 min at room temperature (RT) in the dark. Cells were then washed with 10% FBS in PBS, centrifuged at 391 g for 7 min at RT, and incubated with 10% FBS in PBS for 10 min at 4°C. Surface staining was performed by incubating cells in 100 μL of antibody cocktail (details in **Table S4**) prepared in 10% FBS in PBS for 25 min at 4°C, protected from light. Cells were washed twice with PBS and resuspended in 100 μL MACS buffer (2 mM EDTA (Omega) and 0.5% BSA (MilliporeSigma) in PBS, pH 7.0). Flow cytometry acquisition was performed on a Cytek Aurora (CYTEK Biosciences). Compensation controls were prepared using single-stained UltraComp eBeads (eBiosciences), with 2 μL of each antibody per control. Results were analyzed using FlowJo software (v10.10.0). The gating strategy of PMBC cell frequencies is depicted in **Figure S3B**.

### AIM assay

Cryopreserved PBMCs were thawed at 37°C for 1 min and transferred into 10 mL pre-warmed cell culture medium with 20 μL of Benzonase nuclease (MilliporeSigma). Cell culture medium consisted of RPMI 1640 (Corning, 10-041-CM) supplemented with 5% Human Serum AB (GeminiBio, 110-512), 1% Penicillin:Streptomycin solution (GeminiBio, 400-109), and 1% GlutaMAX (Gibco, 35050061). Cells were centrifuged at 428 g for 5 min at RT and resuspended in cell culture medium for counting and viability assessment using 0.02% Trypan Blue (ThermoFisher) and a hemocytometer. For stimulation assays, 1×10L PBMCs in 100 μL cell culture medium were plated per well in a 96-well plate (GenClone, 25-221) and exposed to one of the five conditions: tetanus toxoid epitope pool (1 μg/mL)(51), aP megapool (1 μg/mL, Bp(E)VAC)(21), non-aP Bp megapool (1 μg/mL, Bp(E)R_EXT)(20), DMSO (0.3%, negative control, MilliporeSigma, D2650), and PHA-L (1 μg/mL, positive control, MilliporeSigma, 431784). Before incubation, antibodies against CD40, CXCR5, and CCR7 (**Table S4**) were added to each well. Cultures were incubated for 20 h at 37°C with 5% CO_2_, followed by the addition of BD GolgiPlug and BD GolgiStop (both 1:1000) which were incubated for 4 h. After incubation, cells were washed once with PBS (Gibco) and stained with 1% Zombie UV fixable viability dye (BioLegend) in PBS and incubated for 15 min at room temperature (RT) in the dark. Cells were then washed with 10% FBS in PBS. Surface staining was performed in 100 μL of antibody/dye cocktail (**Table S4**) for 30 min at 4 °C in the dark. After surface staining, cells were washed twice with 10% FBS in PBS and fixed using Cyto-Fast Fix/Perm Buffer (BioLegend). After fixation, cells were washed using 1x Cyto-Fast Perm wash solution (BioLegend). Then the blocking buffer (10% Human serum in 1x Perm/Wash) was added to each well and incubated for 15 minutes in the dark at RT, followed by intracellular cytokine staining and 30 minutes incubation at RT. Stained cells were centrifuged at 391 g for 5 min at 4°C, washed twice with 1x Cyto-Fast Perm wash solution, and resuspended in MACS buffer (2 mM EDTA, (Omega) and 0.5% BSA (MilliporeSigma) in PBS, pH 7.0). For compensation, beads (Thermo Fisher, 01-2222-42) were washed twice with PBS, centrifuged at 391 g for 2 min at RT, and single-stained with 2 μL of each antibody. Data acquisition was performed on a Cytek Aurora (CYTEK Biosciences) flow cytometer, and analyses were conducted using FlowJo v10.10.0. The AIM gating strategy is shown in **Figure S4B**. Background subtraction was performed by subtracting DMSO control values from megapool responses. The median DMSO frequency of 0.0016% was used as the LLQ, which was added to all measurements before post-/pre-challenge calculations to avoid artificially inflated values near or below the detection limit. For TT correction, measurements were divided by the corresponding TT value for each sample. For DMSO correction, the DMSO value was subtracted from each sample. For percentage-based plots, any post-subtraction values below the LLQ were set to LLQ.

### Fluorospot

Two million PBMCs were plated in 500 μL cell culture medium (formulation described above). Cells were stimulated with 2 μg/mL aP megapool (Bp(E)VAC)(21) or non-aP Bp megapool (Bp(E)R_EXT)(20) for 14 days at 37°C and 5% CO_2_. On day 4, cultures received 1 mL cell culture medium with IL-2 (10 U/mL, Prospec CYT-209B), and on days 7 and 11, 1 mL medium was replaced with fresh medium. Fluorospot PVDF plates (Mabtech, 3654-FL-10) were washed with 70% methanol (MilliporeSigma, 154903), rinsed three times with sterile dH_2_O, and coated overnight at 4°C with a capture antibody mixture containing 10 µg/mL mouse anti-human IFN-γ (Mabtech, clone: 1-D1K), IL-5 (Mabtech, clone: TRFK5), and IL-17 (Mabtech, clone: MT44.6) in PBS (Gibco). The following day, plates were washed three times with PBS and blocked for 1 h at 37°C using cell culture medium. The blocking medium was then replaced with medium containing either 2 µg/mL aP megapool, non-aP Bp megapool, 0.2% DMSO (negative control; MilliporeSigma, D2650), or 20 µg/mL PHA-L (positive control; MilliporeSigma, 431784). After the 14-day culture period, PBMCs were harvested, washed, and enumerated using a hemocytometer. Half a million cells were added to each well containing simulants in a 1:1 ratio and cultured for 24 h at 37°C and 5% CO_2_. Plates were washed five times with 0.05% Tween-20 (MilliporeSigma, PPB005) in PBS and then incubated with the following detection antibodies (Mabtech): anti-IFN-γ 7-B6-1-BAM (1:200), anti-IL-5 5A10-WASP (1:200), and 2 μg/mL anti-IL-17 MT504 biotinylated in 0.1% bovine serum albumin (BSA, MilliporeSigma, A3294) in PBS. Plates were incubated for 2 h at RT, washed five times with 0.05% Tween-20 in PBS, and incubated with the following fluorophore-conjugated detection antibodies (Mabtech): anti-BAM-490, anti-WASP-640, and SA-550, all at 1:200 in 0.1% BSA in PBS, for 1 h at RT in the dark. Plates were washed five times with 0.05% Tween-20 in PBS and fluorescence enhancer (Mabtech, 3641-F10) was applied for 15 min at RT. Plates were air-dried and quantified on a Mabtech IRIS ELISpot/Fluorospot reader. A cytokine response was considered positive when the following three criteria were met: (1) eliciting ≥20 spot-forming cells (SFC) per 10L PBMC after DMSO control subtraction, (2) statistical significance (P ≤ 0.05) by Student’s t-test or by the Poisson distribution test when comparing triplicates with the DMSO control, and (3) a ≥2-fold increase compared to DMSO control. For positive responses, the SFC/10L PBMC was calculated as the mean of triplicate values minus the DMSO background. If the criteria were not met, the value was set to 20.

### Plasma cytokine concentrations

Plasma samples were randomly assigned to 96-well plates for the absolute quantification of 45 different cytokines. The Olink® Target 48 Cytokine panel was run by the Human Immune Monitoring Center of Stanford University (CA, USA). Cytokine measurements were made using the Proximity Extension Assay (PEA) technology(52). Briefly, plasma was incubated with oligonucleotide-conjugated antibodies targeting a cytokine of interest. When both antibodies bound the same antigen, their oligonucleotides came into proximity, allowing hybridization and subsequent qPCR amplification for detection. The resulting cycle threshold (Ct) values were converted to Normalized Protein eXpression (NPX) units, which provide relative protein abundance in plasma. For each cytokine, a standard curve was generated to convert NPX values to absolute concentrations (pg/mL) and to correct for inter-plate batch effects. Additional procedural details are available in the Target 48 User Manual (Olink®, v13). Statistical analyses were conducted in GraphPad Prism (v10.4.2) using Dunnett’s multiple comparisons test within a mixed-effects model to evaluate post- versus pre-challenge differences. Before post-challenge/pre-challenge calculations were made, the 5th percentile value was computed and added to all measurements for each cytokine to avoid inflated fold-change estimates from values near or below the detection limit.

### NPS collection and bulk RNA-Seq

NPS from CHIM participants were collected using Flexible Mini Tip FLOQSwabs with 100mm Breakpoint (COPAN) and placed into 1.5 mL of cold TRIzol reagent. Tubes were vortexed 10 x 3 s at high speed, and this step was repeated after the samples rested for 5 min at RT. Swabs were removed from the tubes, and as much fluid as possible was recovered. Cells in TRIzol were transferred to cryovials, snap-frozen using liquid nitrogen and stored at −80°C until further processing.

NPS were also collected from healthy, non-CHIM participants to evaluate our RNA-Seq deconvolution efforts. These NPS were collected using Puritan Hydraflock swabs (VWR) and vortexed with 10 brief pulses at medium-high speed to release cells. A 70-µm cell strainer (Falcon) was pre-moistened with 1 mL medium (RPMI with 1% P/S and 10% FBS), cells were strained, and swab was rinsed. NPS cells were centrifuged at ∼500 RCF for 6 min at 4°C. A PBMC sample was processed simultaneously to support the gating of immune subsets that are not abundant in NPS. Cells were washed with PBS and centrifuged again with the same settings. Dead cells were stained with Zombie UV L/D in PBS with 1/100 Fc block and incubated for 15 min at RT in a 96-well plate protected from light. Then, cells were washed with PBS with 10% FBS and spun again. Supernatant was removed and cells were incubated with an antibody mixture (**Figure S4**) for 30 min in a dark 37°C incubator. Cells were washed twice with PBS and resuspended in MACS buffer. Cells were kept on ice until measured using a Cytek® Aurora system and subsequently all recollected in RPMI with 20% FCS. Gating strategy is depicted in **Figure S6C-D**. Recollected cells were centrifuged at 500 RCF for 7 min at 4°C. Then, 750 μL TRIzol was added to the cell pellet and samples were stored at −80°C until RNA isolation.

RNA was extracted with on-column DNase treatment using the miRNeasy Mini Kit (Qiagen) following the manufacturer’s instructions. RNA was quantified by qPCR using the housekeeping gene B2M. For cDNA synthesis, 10 ng of RNA was reverse-transcribed and pre-amplified (14 cycles) according to the Smart-Seq2 protocol. cDNA was purified with AMPure XP beads (Beckman Coulter), and 0.5 ng was used for Nextera XT sequencing library preparation with 8-Bp Unique Dual Index (UDI) barcodes (IDT, Illumina) and the Nextera XT DNA Library Preparation Kit (Illumina). To minimize inter-assay variability, all purification steps were performed using an EpMotion 5075 (Eppendorf), and sequencing libraries were prepared in a 96-well format using a Mantis microfluidics liquid dispenser (Formulamatrix). Quality control included assessment of total RNA quality and quantity, optimization of pre-amplification cycle numbers, and fragment size analysis by capillary electrophoresis (5300 Fragment Analyzer, Agilent). cDNA and final libraries were quantified with the Quant-iT PicoGreen assay (Thermo Fisher Scientific). All samples passed quality thresholds, with >70% of fragments ranging from 300–700 base pairs and <5% primer dimers. Libraries were quantified by qPCR using the KAPA Library Quantification Kit (Roche), normalized to equimolar concentrations, and loaded onto a NovaSeq X Plus sequencing platform (Illumina) with a 25B flow cell. Paired-end sequencing (150 base pairs) yielded 4.2 billion total reads, corresponding to an average depth of ∼15 million reads per sample.

### PBMC bulk RNA-Seq

PBMCs (500K) were stored in 750 μL TRIzol at −80°C until further processing. RNA was extracted with on-column DNase treatment using the miRNeasy Mini Kit (Qiagen) following the manufacturer’s instructions. Quantabio Sparq mRNA-Seq kit was used to prepare stranded libraries using 8 ng of high-quality total RNA as input, except in 10 samples where the yield was below 8 ng. Libraries underwent 16-18 PCR amplification cycles and libraries were indexed with xGen ten-nucleotide Unique Dual Indexes (UDIs). Paired-end (2x50 base pairs) sequencing was performed on a NovaSeq 6000 targeting 40 million reads per library.

### Bioinformatics NPS and PBMC RNA-Seq

Paired-end reads that passed Illumina quality filters were further screened to remove sequences aligning to tRNA, rRNA, adapter sequences, and spike-in controls. The remaining reads were aligned to the GRCh38 human reference genome with Gencode v27 annotations using STAR (v2.6.1)(53). Read complexity was assessed with PRINSEQ Lite (v0.20.3) (54), and low-complexity reads (DUST score > 4) were excluded from the BAM files. The alignment results were further processed using SAMtools (55). Gene-level counts were then obtained using featureCounts (v1.6.5)(56) using the default option along with a minimum quality cut-off (Phred > 10), and transcripts per million (TPM) reads values were calculated. Using HUGO Gene Nomenclature Committee annotations, T cell receptor (TCR) transcripts were aggregated into four groups: TCR-α, TCR-β, TCR-γ, and TCR-δ. B cell receptor (BCR) transcripts were aggregated into light chains (*IGK* and *IGL*) and heavy chains (*IGH*-), with isotype transcripts (*IGHG1-4, IGHM, IGHD, IGHE, IGHA1-2*) analyzed separately. Transcripts were filtered using EdgeR’s *filterByExpr* function (v4.2.2)(57) with “participants + timepoint” as the design. For each participant, baseline TPMs were averaged and raw counts were summed. Differential expression was assessed using the DESeq2 package (v1.44.0)(58) for NPS and EdgeR for PBMCs in an R (v4.4.0) environment, with gene expression called differential with an FDR<0.05. Gene set enrichment analysis (GSEA) was performed using Metascape(59) and GSEA (v4.4.0)(60), both with the Gene Ontology (GO) gene sets(61).

### NPS deconvolution

#### Pre-processing of single-cell datasets

For cell-type-specific marker selection, single-cell RNA-Seq raw counts were extracted from publicly available datasets of the upper airway in COVID-19 patients and healthy controls(26, 62). TCR and BCR transcripts were aggregated as described above. Cell-type annotations identified in the publications were matched to the study’s target cell types (**Table S5**). The effect of combining CD4+ and CD8+ T cells was also evaluated. To refine cell-type resolution in the Ziegler *et al.* dataset, CellTypist with the “Immune_All_High” reference(63, 64) was applied, and resulting subtypes were mapped to major populations (**Table S5**).

#### Marker extraction from single-cell datasets to create signature matrices

Cell-type-specific markers were then identified using the Seurat v5 *FindAllMarkers()* function with the Wilcoxon test. Genes were retained if they met thresholds of FDR < 0.05, log2 fold change > 2.5, and normalized expression > –12 (using the BayesPrism normalisation function(65). After being ranked by specificity (calculated as the highest expression in a cell type over the total expression in all cell types), the top 150 genes per cell type were selected to construct signature matrices for deconvolution. Counts were aggregated per cell type and normalized as counts per million (CPM). For methods that generate their own signature matrices, single-cell count matrices were downsampled to 5000 cells per cell type. These matrices, along with their cell type annotations, were passed to CIBERSORTx and DWLS to produce internal signature matrices.

#### Benchmarking

NPS bulk RNA-Seq data and flow cytometry–derived cell-type frequencies from the same non-CHIM participants were used for benchmarking. Ensembl IDs were converted to gene symbols using GRCh38, and duplicate entries were resolved by retaining the transcript with the highest expression. This RNA-Seq matrix was used in conjunction with the generated signature matrices to estimate the cell type proportions of the non-CHIM participant samples. Estimated fractions from each deconvolution method were compared to flow cytometry–derived cell-type frequencies by Pearson correlation and root mean square error (RMSE). We tested three methods (described below) with the signature matrices described above. The deconvolution–signature combination (CIBERSORTx with Ramirez signature) yielded the highest correlation and lowest RMSE was then applied to CHIM NPS RNA-Seq data. NPS samples predicted to contain <1% CD45^−^ cells were excluded from downstream analysis.

#### Deconvolution methods for frequency deconvolution

Deconvolution was performed using multiple algorithms. FARDEEP(66) was applied to CPM-normalized bulk RNA-Seq with default parameters, except that the intercept was set to FALSE and α1 was tuned (0.4 for the Ziegler–Celltypist signature, and 1.0 for all other signatures). CIBERSORTx(25) was applied in relative mode with batch correction (which rescaled the bulk RNA-Seq data according to the distribution observed in the signature matrix); default settings were used for internally generated signatures. DWLS(67) was applied to raw counts using default parameters for both custom and internal signature matrices.

#### GEP deconvolution

Gene expression profile (GEP) deconvolution was performed on the CHIM samples using CIBERSORTx. High-resolution deconvolution generated cell-type-specific GEPs for each sample, enabling comparison of gene expression between outcome groups and timepoints within individual cell types. Batch correction was applied during deconvolution using default parameters. Among the 1196 DEGs observed on day 7 in symptomatic participants from bulk RNA-Seq data, we took 1,000 genes for high-resolution imputation (the maximum allowed by CIBERSORTx): all 46 downregulated genes, plus the top 954 up-regulated genes based on log fold change. Differential expression within each subset was assessed using the EdgeR with gene expression called differential with a P<0.05.

## Data availability

PBMC and NPS gene expression data are deposited in the Gene Expression Omnibus (GEO) under the GEO SuperSeries accession number GSE310241 (reviewer token: xxx). Fastq files are stored in the Sequence Read Archive (SRA) under the BioProject ID PRJNA1354793.

## Acknowledgements

This work was supported by CDC (75D30122C15481).

- This publication includes data generated at the UC San Diego IGM Genomics Center utilizing an Illumina X Plus that was purchased with funding from a National Institutes of Health SIG grant (#S10 OD026929), and the authors would like to thank Gregory Seumois, Monalisa Mondal, and Veronica Tapia Guillen for their contributions.
- The authors would like to thank all study participants and the La Jolla Institute for Immunology Bioinformatics (J. Greenbaum) and Next Generation Sequencing (S. Alarcón; RRID:SCR_023107) core facilities for their contribution. The NovaSeq 6000 was acquired through the Shared Instrumentation Grant (SIG) Program: S10OD025052. Additionally, we would like to acknowledge the Human Immune Monitoring Center of Stanford University for their Olink services and the PERISCOPE consortium (funded by the European Commission, Innovative Medicines Initiative 2, Grant No 115910) members for kindly providing the serum IgG titers of their phase A study.
- Figure 1A-B and S6C were created using BioRender.com [license links].

## Author contributions

Study design: B.P., S.H., M.E., L.W.; Data collection: Z.R., J.L., L.W.; Data analysis and graphical representation: L.W.; RNA-Seq deconvolution: Z.R., N.T., A.G., L.W.; Data and\ specimen management: P.S., J.S., A.A., A.S., S.O.; Project management: S.O., M.K., A.F.; Manuscript writing and feedback: L.W., B.P., S.H. P.S., N.T., S.O.; Study initiation and supervision: B.P.

## Disclaimer

The findings and conclusions in this report are those of the authors and do not necessarily represent the official position of the Centers for Disease Control and Prevention, US Department of Health and Human Services.

## Supplemental information

**Figure S1.**
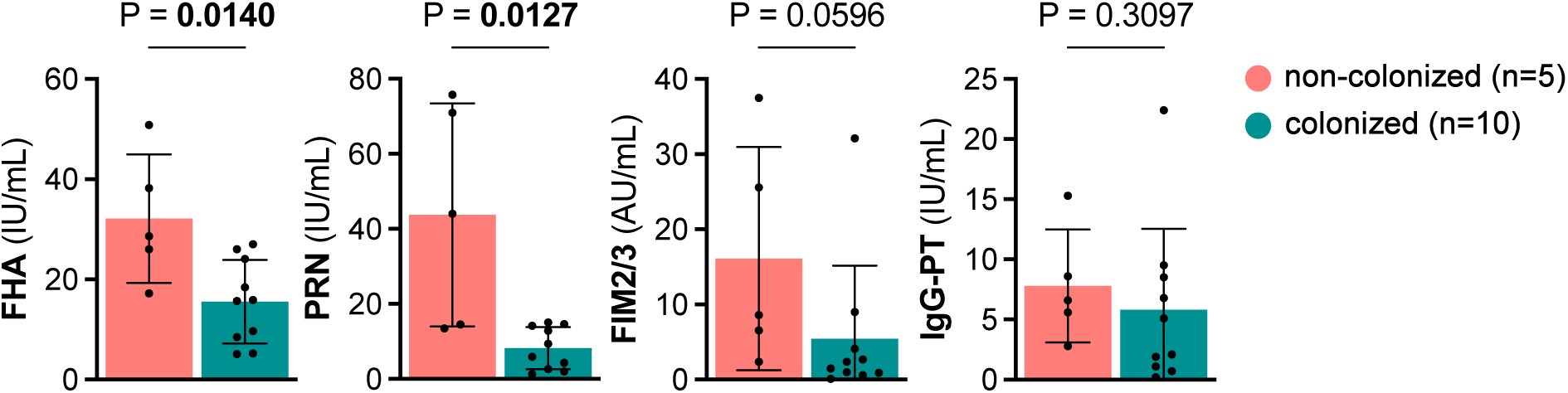
Non-colonized (non-infected) participants from the PERISCOPE Bp CHIM study had higher baseline Bp-specific IgG titers. Serum IgG titers against Bp antigens (PT, PRN, FHA, FIM) before Bp inoculation per outcome definition. Mean bars with SD are depicted per outcome definition. P values were calculated by performing Mann-Whitney tests. Raw data were derived from the PERISCOPE Bp CHIM study.

**Figure S2.**
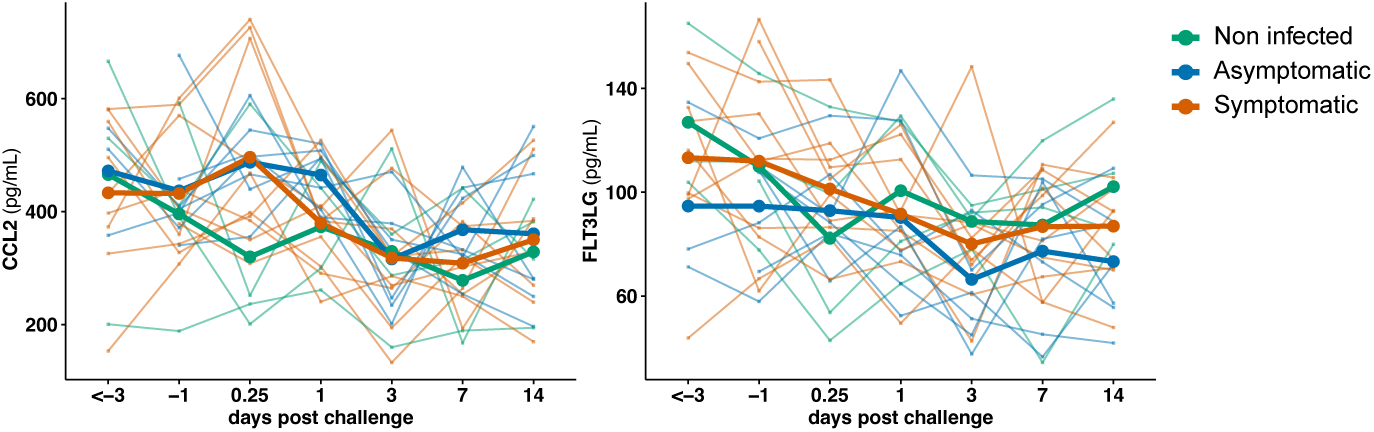
Bp challenge decreased systemic mediators independent of clinical outcome. Plasma CCL2 and FLT3LG concentrations (pg/mL) over time and coloured per outcome definition. Mean lines are shown per outcome group.

**Figure S3.**
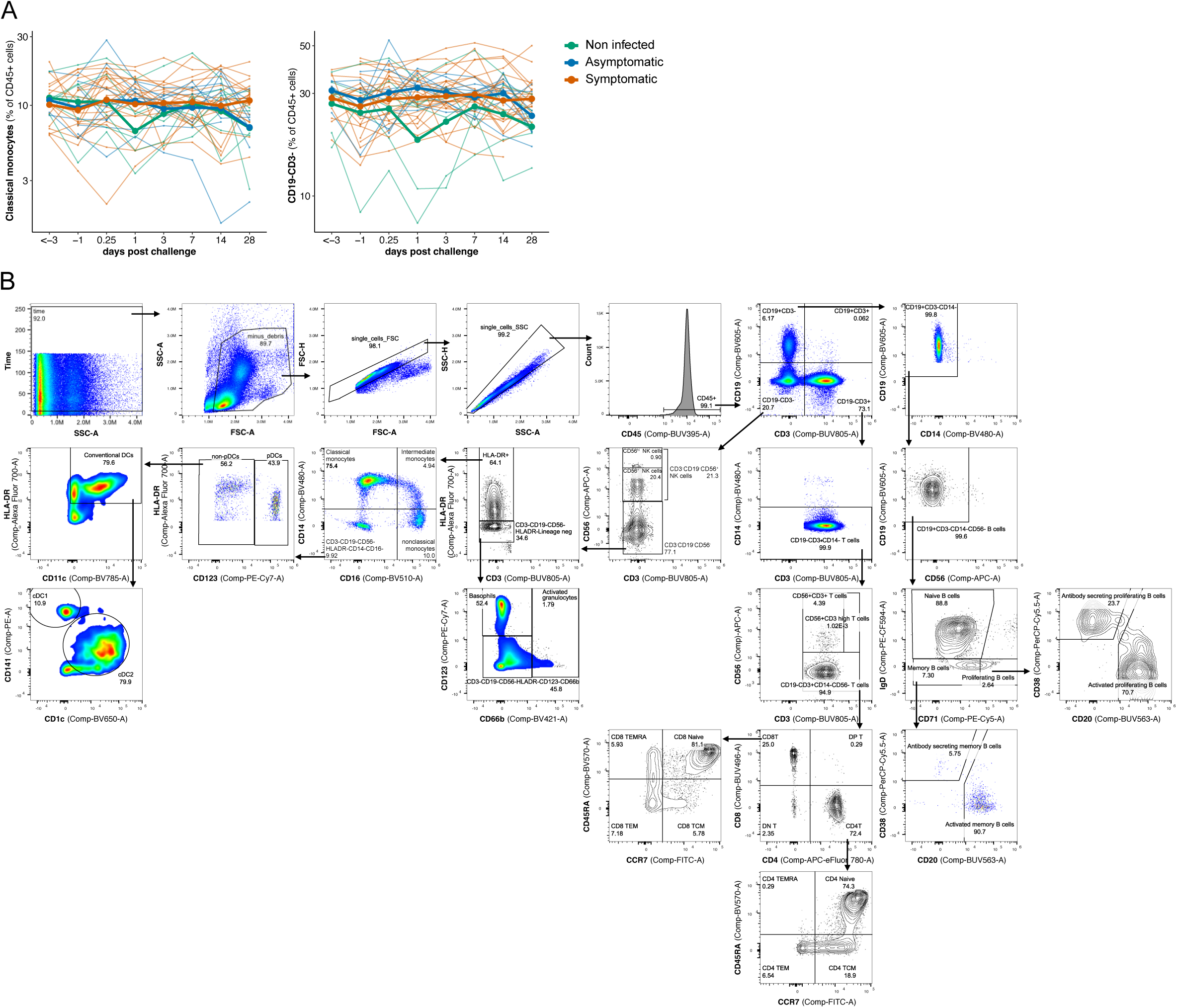
Dynamic and clinically associated changes in blood immune cell frequencies following Bp challenge. **A)** PBMC subset frequencies of CD19-CD3- (left) and classical monocytes (right) over time. Mean lines are depicted per outcome definition. **B)** Flow cytometry gating strategy of PBMC immune cell subsets.

**Figure S4.**
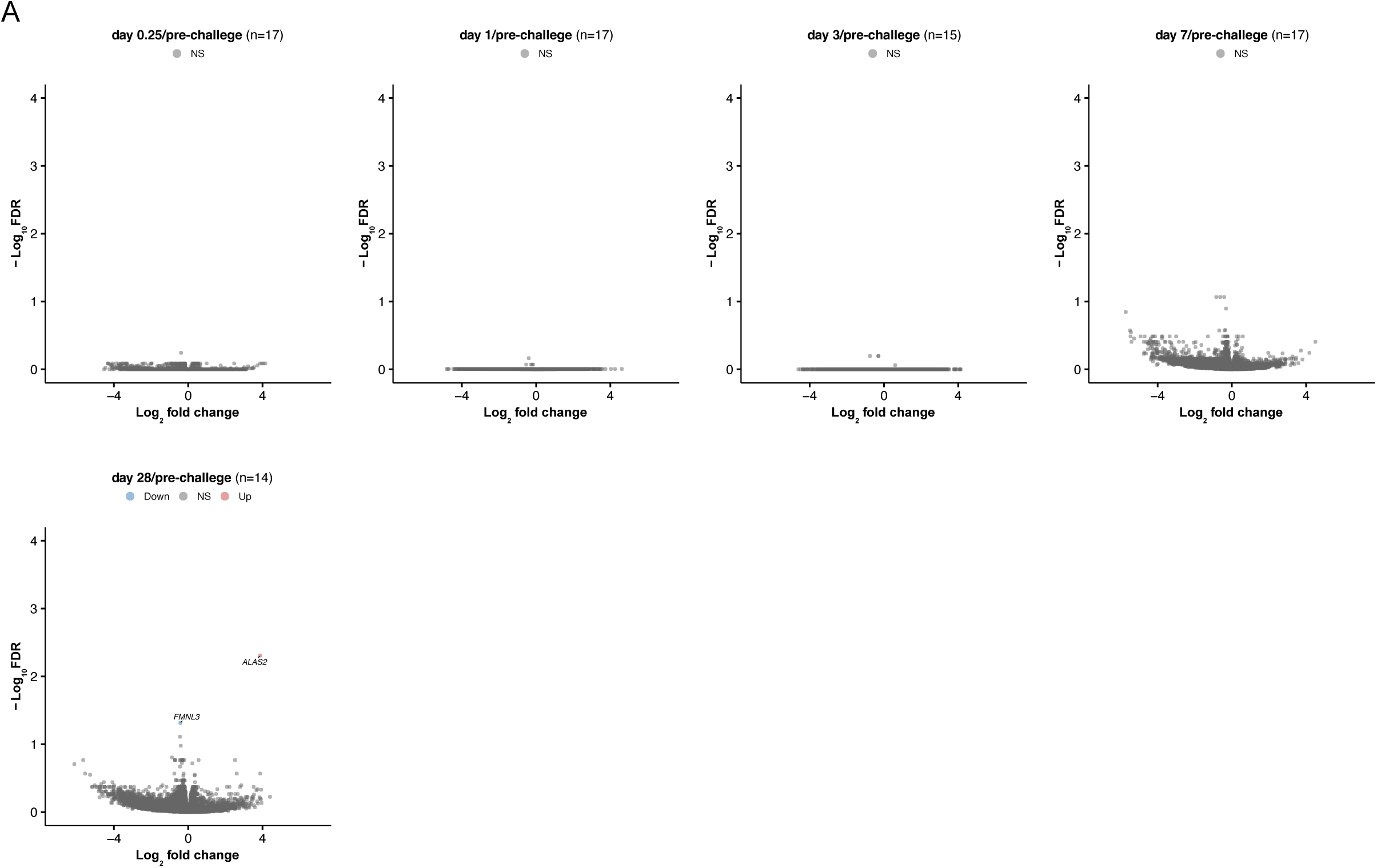
Limited transcriptional changes in PBMCs following Bp challenge. Volcano plots showing the log_2_(fold change) and −log_10_(FDR) of the DEG in PBMCs of symptomatic people on day 0.25, 1, 3, 7, and 28 vs. pre-challenge with downregulated genes in blue and upregulated genes in red (FDR < 0.05).

**Figure S5.**
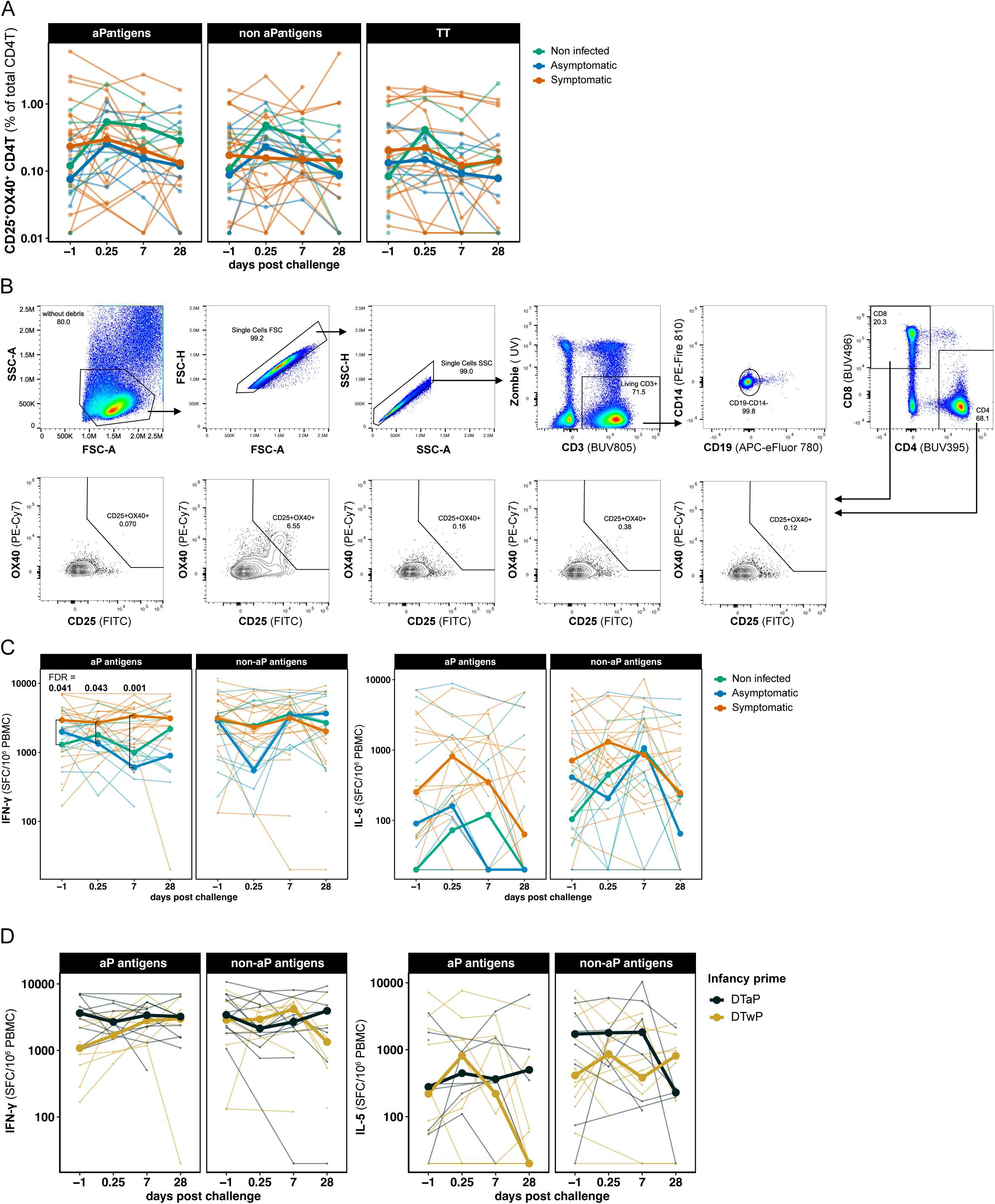
Bp exposure and infection did not affect antigen-specific T cell activation and polarization in the blood. **A)** Activation-induced marker (AIM) assays: PBMCs were stimulated with 1 μg/mL of either the pertussis (containing aP or non-aP Bp vaccine antigens) or tetanus peptide megapools for 18–24 h. CD25^+^OX40^+^ CD4L T cells were measured by flow cytometry and expressed as a percentage of total CD4L T cells, normalized to DMSO responses. Mean lines are depicted by clinical outcome; P values were calculated by multiple two-tailed Wilcoxon matched-pairs tests. **B)** Flow cytometry gating strategy for CD25^+^OX40^+^ CD4^+^ or CD8^+^ T cells. **C-D)** IFN-γ or IL-5 producing cells (spot-forming cells, SFC) were measured by Fluorospot following 14-day stimulation with aP or non-aP Bp megapools. Median lines are depicted by C) outcome definition and D) primary pertussis vaccine group within symptomatic participants. P values were calculated using multiple two-tailed Mann-Whitney tests.

**Figure S6.**
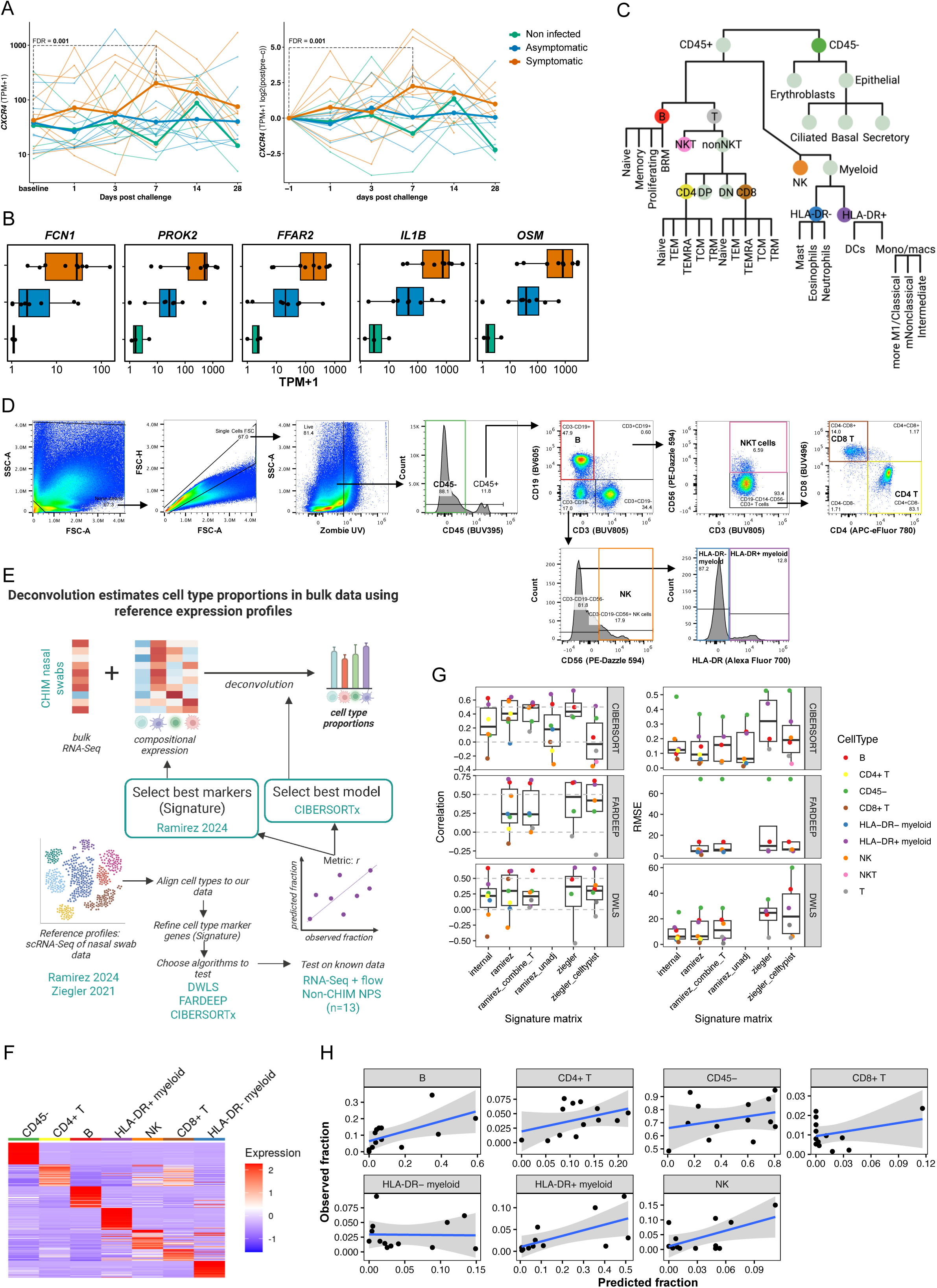
Bp challenge induced a nasal inflammatory response on day 7, specifically in people who later became symptomatic. **A)** *CXCR4* gene expression (TPM+1) over time per outcome definition (left) and normalized to baseline expression (right). **B)** *FFAR2, FCN1, OSM, IL1B,* and *PROK2* gene expression (TPM+1) on day 7 post-challenge per outcome definition. **C)** Schematic representation of the NPS cell subsets and their hierarchical relationships identified by flow cytometry, created using BioRender [license link]. **D)** Flow cytometry gating strategy of NPS cell subsets. **E)** Transcriptomic deconvolution strategy to estimate NPS cell frequencies. Bottom left: cell type-specific markers were identified and used to build signature matrices. Three deconvolution tools: DWLS, FARDEEP, CIBERSORTx were tested. To benchmark the different methods and signature matrices, RNAseq and cell type frequency data were generated for non-CHIM NPS (bottom right). The final model was used to estimate cell type proportions in the Bp-challenged participants. **F)** Specificity of markers used for deconvolution. Heatmap shows scaled expression of the selected markers (derived from Ramirez et. al. 2024) with red and blue indicating higher and lower expression respectively. **G)** Boxplots show correlations (left) and root mean square error (RMSE; right) between observed and predicted cell type fractions estimated with different deconvolution tools (facets) and signature matrices (x-axis). H) Correlations of observed vs predicted proportions in non-CHIM NPS. Each point represents a sample. The blue line represents a fitted regression line, with confidence intervals represented in grey.

## References

1. Smout E, Mellon D, Rae M. Whooping cough rises sharply in UK and Europe. BMJ. 2024;385:q736.

2. (CDC) CfDCaP. Pertussis Surveillance and Trends 2025 [updated December 2, 2025. Available from: https://www.cdc.gov/pertussis/php/surveillance/index.html.

3. Domenech de Celles M, Rohani P. Pertussis vaccines, epidemiology and evolution. Nat Rev Microbiol. 2024;22(11):722–35.

4. Ross PJ, Sutton CE, Higgins S, Allen AC, Walsh K, Misiak A, et al. Relative contribution of Th1 and Th17 cells in adaptive immunity to Bordetella pertussis: towards the rational design of an improved acellular pertussis vaccine. PLoS Pathog. 2013;9(4):e1003264.

5. Mahon Bp, Sheahan BJ, Griffin F, Murphy G, Mills KH. Atypical disease after Bordetella pertussis respiratory infection of mice with targeted disruptions of interferon-gamma receptor or immunoglobulin mu chain genes. J Exp Med. 1997;186(11):1843–51.

6. Warfel JM, Zimmerman LI, Merkel TJ. Acellular pertussis vaccines protect against disease but fail to prevent infection and transmission in a nonhuman primate model. Proc Natl Acad Sci U S A. 2014;111(2):787–92.

7. Witt MA, Arias L, Katz PH, Truong ET, Witt DJ. Reduced risk of pertussis among persons ever vaccinated with whole cell pertussis vaccine compared to recipients of acellular pertussis vaccines in a large US cohort. Clin Infect Dis. 2013;56(9):1248–54.

8. Klein NP, Bartlett J, Fireman B, Rowhani-Rahbar A, Baxter R. Comparative effectiveness of acellular versus whole-cell pertussis vaccines in teenagers. Pediatrics. 2013;131(6):e1716–22.

9. Higgs R, Higgins SC, Ross PJ, Mills KH. Immunity to the respiratory pathogen Bordetella pertussis. Mucosal Immunol. 2012;5(5):485–500.

10. Valeri V, Sochon A, Cousu C, Chappert P, Lecoeuche D, Blanc P, et al. The whole-cell pertussis vaccine imposes a broad effector B cell response in mouse heterologous prime-boost settings. JCI Insight. 2022;7(21).

11. Ryan M, Murphy G, Gothefors L, Nilsson L, Storsaeter J, Mills KH. Bordetella pertussis respiratory infection in children is associated with preferential activation of type 1 T helper cells. J Infect Dis. 1997;175(5):1246–50.

12. Ausiello CM, Urbani F, la Sala A, Lande R, Cassone A. Vaccine- and antigen-dependent type 1 and type 2 cytokine induction after primary vaccination of infants with whole-cell or acellular pertussis vaccines. Infect Immun. 1997;65(6):2168–74.

13. Weaver KL, Bitzer GJ, Wolf MA, Pyles GM, DeJong MA, Dublin SR, et al. Intranasal challenge with B. pertussis leads to more severe disease manifestations in mice than aerosol challenge. PLoS One. 2023;18(11):e0286925.

14. Brotons P, de Paz HD, Toledo D, Villanova M, Plans P, Jordan I, et al. Differences in Bordetella pertussis DNA load according to clinical and epidemiological characteristics of patients with whooping cough. J Infect. 2016;72(4):460–7.

15. ElSherif MS, Redden KL, Langley JM, Ye L, Blanchard W, Smith B, et al. A controlled human infection model for symptomatic pertussis in North America using the pertactin-producing clinical isolate D420. medRxiv. 2026(TBD).

16. de Graaf H, Ibrahim M, Hill AR, Gbesemete D, Vaughan AT, Gorringe A, et al. Controlled Human Infection With Bordetella pertussis Induces Asymptomatic, Immunizing Colonization. Clin Infect Dis. 2020;71(2):403–11.

17. Diks AM, de Graaf H, Teodosio C, Groenland RJ, de Mooij B, Ibrahim M, et al. Distinct early cellular kinetics in participants protected against colonization upon Bordetella pertussis challenge. J Clin Invest. 2023;133(5).

18. Willemsen L, Lee J, Shinde P, Soldevila F, Aoki M, Orfield S, et al. Th1 polarization in Bordetella pertussis vaccine responses is maintained through a positive feedback loop. Nat Commun. 2025;16(1):3132.

19. Dan JM, Lindestam Arlehamn CS, Weiskopf D, da Silva Antunes R, Havenar-Daughton C, Reiss SM, et al. A Cytokine-Independent Approach To Identify Antigen-Specific Human Germinal Center T Follicular Helper Cells and Rare Antigen-Specific CD4+ T Cells in Blood. J Immunol. 2016;197(3):983–93.

20. da Silva Antunes R, Garrigan E, Quiambao LG, Dhanda SK, Marrama D, Westernberg L, et al. T cell reactivity to Bordetella pertussis is highly diverse regardless of childhood vaccination. Cell Host Microbe. 2023;31(8):1404–16 e4.

21. da Silva Antunes R, Babor M, Carpenter C, Khalil N, Cortese M, Mentzer AJ, et al. Th1/Th17 polarization persists following whole-cell pertussis vaccination despite repeated acellular boosters. J Clin Invest. 2018;128(9):3853–65.

22. Bancroft T, Dillon MB, da Silva Antunes R, Paul S, Peters B, Crotty S, et al. Th1 versus Th2 T cell polarization by whole-cell and acellular childhood pertussis vaccines persists upon re-immunization in adolescence and adulthood. Cell Immunol. 2016;304-305:35–43.

23. van der Lee S, Hendrikx LH, Sanders EAM, Berbers GAM, Buisman AM. Whole-Cell or Acellular Pertussis Primary Immunizations in Infancy Determines Adolescent Cellular Immune Profiles. Front Immunol. 2018;9:51.

24. da Silva Antunes R, Quiambao LG, Sutherland A, Soldevila F, Dhanda SK, Armstrong SK, et al. Development and Validation of a Bordetella pertussis Whole-Genome Screening Strategy. J Immunol Res. 2020;2020:8202067.

25. Newman AM, Steen CB, Liu CL, Gentles AJ, Chaudhuri AA, Scherer F, et al. Determining cell type abundance and expression from bulk tissues with digital cytometry. Nat Biotechnol. 2019;37(7):773–82.

26. Ramirez SI, Faraji F, Hills LB, Lopez PG, Goodwin B, Stacey HD, et al. Immunological memory diversity in the human upper airway. Nature. 2024;632(8025):630–6.

27. Bond M, Chase AJ, Baker AH, Newby AC. Inhibition of transcription factor NF-kappaB reduces matrix metalloproteinase-1, -3 and -9 production by vascular smooth muscle cells. Cardiovasc Res. 2001;50(3):556–65.

28. Sun R, Hedl M, Abraham C. TNFSF15 Promotes Antimicrobial Pathways in Human Macrophages and These Are Modulated by TNFSF15 Disease-Risk Variants. Cell Mol Gastroenterol Hepatol. 2021;11(1):249–72.

29. Fang L, Che Y, Zhang C, Huang J, Lei Y, Lu Z, et al. PLAU directs conversion of fibroblasts to inflammatory cancer-associated fibroblasts, promoting esophageal squamous cell carcinoma progression via uPAR/Akt/NF-kappaB/IL8 pathway. Cell Death Discov. 2021;7(1):32.

30. Murphy EP, Crean D. Molecular Interactions between NR4A Orphan Nuclear Receptors and NF-kappaB Are Required for Appropriate Inflammatory Responses and Immune Cell Homeostasis. Biomolecules. 2015;5(3):1302–18.

31. Wamsley JJ, Kumar M, Allison DF, Clift SH, Holzknecht CM, Szymura SJ, et al. Activin upregulation by NF-kappaB is required to maintain mesenchymal features of cancer stem-like cells in non-small cell lung cancer. Cancer Res. 2015;75(2):426–35.

32. Browatzki M, Schmidt J, Kubler W, Kranzhofer R. Endothelin-1 induces interleukin-6 release via activation of the transcription factor NF-kappaB in human vascular smooth muscle cells. Basic Res Cardiol. 2000;95(2):98–105.

33. Storsaeter J, Hallander HO, Gustafsson L, Olin P. Levels of anti-pertussis antibodies related to protection after household exposure to Bordetella pertussis. Vaccine. 1998;16(20):1907–16.

34. Deen JL, Mink CA, Cherry JD, Christenson PD, Pineda EF, Lewis K, et al. Household contact study of Bordetella pertussis infections. Clin Infect Dis. 1995;21(5):1211–9.

35. de Graaf H, Gbesemete DF, Hill AR, Froberg J, Ibrahim MM, Dale AP, et al. Safety, colonisation kinetics, transmissibility, and immune correlates of protection in healthy adults inoculated with Bordetella pertussis in England: a single-centre, open-label, phase 1, controlled human infection study. Lancet Microbe. 2026:101313.

36. Cherry JD, Gornbein J, Heininger U, Stehr K. A search for serologic correlates of immunity to Bordetella pertussis cough illnesses. Vaccine. 1998;16(20):1901–6.

37. Barkoff AM, Grondahl-Yli-Hannuksela K, He Q. Seroprevalence studies of pertussis: what have we learned from different immunized populations. Pathog Dis. 2015;73(7).

38. Long SS, Welkon CJ, Clark JL. Widespread silent transmission of pertussis in families: antibody correlates of infection and symptomatology. J Infect Dis. 1990;161(3):480–6.

39. Althouse BM, Scarpino SV. Asymptomatic transmission and the resurgence of Bordetella pertussis. BMC Med. 2015;13:146.

40. Craig R, Kunkel E, Crowcroft NS, Fitzpatrick MC, de Melker H, Althouse BM, et al. Asymptomatic Infection and Transmission of Pertussis in Households: A Systematic Review. Clin Infect Dis. 2020;70(1):152–61.

41. Kristiansen PA, Ba AK, Ouedraogo AS, Sanou I, Ouedraogo R, Sangare L, et al. Persistent low carriage of serogroup A Neisseria meningitidis two years after mass vaccination with the meningococcal conjugate vaccine, MenAfriVac. BMC Infect Dis. 2014;14:663.

42. Stephens DS. Protecting the herd: the remarkable effectiveness of the bacterial meningitis polysaccharide-protein conjugate vaccines in altering transmission dynamics. Trans Am Clin Climatol Assoc. 2011;122:115–23.

43. Wilk MM, Misiak A, McManus RM, Allen AC, Lynch MA, Mills KHG. Lung CD4 Tissue-Resident Memory T Cells Mediate Adaptive Immunity Induced by Previous Infection of Mice with Bordetella pertussis. J Immunol. 2017;199(1):233–43.

44. McCarthy KN, Hone S, McLoughlin RM, Mills KHG. IL-17 and IFN-gamma-producing Respiratory Tissue-Resident Memory CD4 T Cells Persist for Decades in Adults Immunized as Children With Whole-Cell Pertussis Vaccines. J Infect Dis. 2024;230(3):e518–e23.

45. Fedele G, Bianco M, Ausiello CM. The virulence factors of Bordetella pertussis: talented modulators of host immune response. Arch Immunol Ther Exp (Warsz). 2013;61(6):445–57.

46. Munoz Navarrete K, Edwards KM, Mills KHG, Kamanova J, Rodriguez ME, Gorringe A, et al. Highlights of the 14th International Bordetella Symposium. mSphere. 2025;10(6):e0018925.

47. Jazayeri SD, Borkner L, Sutton CE, Mills KHG. Respiratory immunization using antibiotic-inactivated Bordetella pertussis confers T cell-mediated protection against nasal infection in mice. Nat Microbiol. 2025;10(12):3094–106.

48. Donnelly S, Loscher CE, Lynch MA, Mills KH. Whole-cell but not acellular pertussis vaccines induce convulsive activity in mice: evidence of a role for toxin-induced interleukin-1beta in a new murine model for analysis of neuronal side effects of vaccination. Infect Immun. 2001;69(7):4217–23.

49. McGuirk P, Mahon Bp, Griffin F, Mills KH. Compartmentalization of T cell responses following respiratory infection with Bordetella pertussis: hyporesponsiveness of lung T cells is associated with modulated expression of the co-stimulatory molecule CD28. Eur J Immunol. 1998;28(1):153–63.

50. Dunne PJ, Moran B, Cummins RC, Mills KH. CD11c+CD8alpha+ dendritic cells promote protective immunity to respiratory infection with Bordetella pertussis. J Immunol. 2009;183(1):400–10.

51. da Silva Antunes R, Paul S, Sidney J, Weiskopf D, Dan JM, Phillips E, et al. Definition of Human Epitopes Recognized in Tetanus Toxoid and Development of an Assay Strategy to Detect Ex Vivo Tetanus CD4+ T Cell Responses. PLoS One. 2017;12(1):e0169086.

52. Assarsson E, Lundberg M, Holmquist G, Bjorkesten J, Thorsen SB, Ekman D, et al. Homogenous 96-plex PEA immunoassay exhibiting high sensitivity, specificity, and excellent scalability. PLoS One. 2014;9(4):e95192.

53. Dobin A, Davis CA, Schlesinger F, Drenkow J, Zaleski C, Jha S, et al. STAR: ultrafast universal RNA-seq aligner. Bioinformatics. 2013;29(1):15–21.

54. Schmieder R, Edwards R. Quality control and preprocessing of metagenomic datasets. Bioinformatics. 2011;27(6):863–4.

55. Li H, Handsaker B, Wysoker A, Fennell T, Ruan J, Homer N, et al. The Sequence Alignment/Map format and SAMtools. Bioinformatics. 2009;25(16):2078–9.

56. Liao Y, Smyth GK, Shi W. featureCounts: an efficient general purpose program for assigning sequence reads to genomic features. Bioinformatics. 2014;30(7):923–30.

57. Chen Y, Chen L, Lun ATL, Baldoni PL, Smyth GK. edgeR v4: powerful differential analysis of sequencing data with expanded functionality and improved support for small counts and larger datasets. Nucleic Acids Res. 2025;53(2).

58. Love MI, Huber W, Anders S. Moderated estimation of fold change and dispersion for RNA-seq data with DESeq2. Genome Biol. 2014;15(12):550.

59. Zhou Y, Zhou B, Pache L, Chang M, Khodabakhshi AH, Tanaseichuk O, et al. Metascape provides a biologist-oriented resource for the analysis of systems-level datasets. Nat Commun. 2019;10(1):1523.

60. Subramanian A, Tamayo P, Mootha VK, Mukherjee S, Ebert BL, Gillette MA, et al. Gene set enrichment analysis: a knowledge-based approach for interpreting genome-wide expression profiles. Proc Natl Acad Sci U S A. 2005;102(43):15545–50.

61. Gene Ontology C, Aleksander SA, Balhoff J, Carbon S, Cherry JM, Drabkin HJ, et al. The Gene Ontology knowledgebase in 2023. Genetics. 2023;224(1).

62. Ziegler CGK, Miao VN, Owings AH, Navia AW, Tang Y, Bromley JD, et al. Impaired local intrinsic immunity to SARS-CoV-2 infection in severe COVID-19. Cell. 2021;184(18):4713–33 e22.

63. Xu C, Prete M, Webb S, Jardine L, Stewart BJ, Hoo R, et al. Automatic cell-type harmonization and integration across Human Cell Atlas datasets. Cell. 2023;186(26):5876–91 e20.

64. Dominguez Conde C, Xu C, Jarvis LB, Rainbow DB, Wells SB, Gomes T, et al. Cross-tissue immune cell analysis reveals tissue-specific features in humans. Science. 2022;376(6594):eabl5197.

65. Chu T, Wang Z, Pe’er D, Danko CG. Cell type and gene expression deconvolution with BayesPrism enables Bayesian integrative analysis across bulk and single-cell RNA sequencing in oncology. Nat Cancer. 2022;3(4):505–17.

66. Hao Y, Yan M, Heath BR, Lei YL, Xie Y. Fast and robust deconvolution of tumor infiltrating lymphocyte from expression profiles using least trimmed squares. PLoS Comput Biol. 2019;15(5):e1006976.

67. Tsoucas D, Dong R, Chen H, Zhu Q, Guo G, Yuan GC. Accurate estimation of cell-type composition from gene expression data. Nat Commun. 2019;10(1):2975

